# Clinical Implications of Inflammation-Mediated Dysregulation of Glial Cell Fate: A Systematic Review for Early Brain Tumor Risk Detection and Improved Patient Outcomes

**DOI:** 10.1101/2025.08.05.25333055

**Authors:** Ovais Shafi, Muhammad Waqas, Luqman Naseer Virk, Sameed Abdul Hameed Siddiqui, Osama Jawed Khan, Ibrahim Abdul Rahman, Ameerah Karim, Muhammad Danyal Khalid, Maleeha Anum, Sameeta Makwana, Manesh Kumar, Aamir Ameer, Finza Kanwal, FNU Raveena, Javeryah Rafiq Shaikh, Muhammad Danial Yaqub, Ayesha Saeed, FNU Aakash

## Abstract

**Objective:** This study aims to investigate how neuroinflammation alters gliogenic regulatory pathways and glial cell fate decisions, with a focus on identifying clinically relevant mechanisms that may increase susceptibility to glial-derived brain tumors. By linking inflammatory signaling to early disruptions in gliogenesis, this study seeks to inform future strategies for risk assessment, early detection, and potential therapeutic intervention in inflammation-associated neuro-oncology.

**Background:** Neuroinflammation alters gliogenic signaling by disrupting key regulatory pathways such as FGFR3, JAK-STAT, STAT3, and others. This dysregulation affects glial cell differentiation and lineage decisions, contributing to impaired neurodevelopment and increased susceptibility to glial-derived brain tumors, particularly gliomas. Understanding these inflammatory mechanisms is essential for identifying early biomarkers, evaluating long-term tumor risk, and developing strategies to prevent or mitigate neuro-oncological outcomes in individuals with acute or chronic CNS inflammation.

**Methods:** Databases, including PubMed, MEDLINE, Google Scholar, and both open-access and subscription-based journals, were searched without date restrictions to investigate the brain inflammation induced disruption of gliogenic regulators (FGFR3, JAK-STAT, STAT3, S100, Hey1, HES1, DTX, IL-6, NF-κB, Neuregulin-1, MAPK/MEK, E2F/TCFL2, NFIX/Ephrins/Netrins) resulting in glial cell fate dysregulations and Its contribution to brain tumor risk. Studies meeting the criteria outlined in the methods section were systematically reviewed to address the research question. This study adheres to the PRISMA (Preferred Reporting Items for Systematic Reviews and Meta-Analyses) guidelines.

**Results:** Neuroinflammation significantly alters gliogenic regulators, including FGFR3, JAK-STAT, STAT3, S100, IL-6, and NF-κB, disrupting normal glial maturation, migration, and lineage commitment. This leads to aberrant cell fate decisions, sustained proliferation, and genomic instability, conditions favorable for glioma initiation. Pro-inflammatory pathways, notably IL-6 and NF-κB, drive oxidative stress and cellular dysfunction, reinforcing the tumor-permissive environment. These insights point to the clinical relevance of inflammation-driven glial dysregulation in brain tumor pathogenesis.

**Conclusion:** Disruption of gliogenic regulators by neuroinflammation alters glial fate specification, promotes proliferative stress, and contributes to oncogenic transformation within the CNS. These findings emphasize the need to clinically monitor individuals with significant or recurrent brain inflammation for long-term neuro-oncological risks. Early recognition of these disturbances may guide risk stratification, surveillance, and development of anti-inflammatory or gliogenesis-targeted interventions to prevent glial-origin brain tumors.

## Background

The relationship between brain inflammation and the regulation of gliogenesis has become a critical area of research due to its potential implications for both normal brain development and the pathogenesis of brain tumors. Brain inflammation also in conditions like meningitis and encephalitis profoundly impacts glial cells, which are essential for maintaining homeostasis, providing support to neurons, and participating in immune responses within the central nervous system (CNS) [1]. In both diseases, pathogens such as bacteria, viruses, or fungi invade the CNS, leading to an inflammatory response characterized by the activation of microglia, astrocytes, and oligodendrocytes. Microglia, the resident immune cells of the brain, are among the first responders to infection. Upon activation, they undergo morphological changes, shifting from a ramified to an amoeboid form, and release a cascade of pro-inflammatory cytokines like tumor necrosis factor-alpha (TNF-α), interleukin-1β (IL-1β), and interleukin-6 (IL-6). While initially protective, this heightened activation can become detrimental, promoting neurotoxicity and contributing to secondary neuronal injury. Astrocytes also become reactive during meningitis and encephalitis, a process termed astrogliosis. Reactive astrocytes upregulate the expression of glial fibrillary acidic protein (GFAP), hypertrophy, and secrete various inflammatory mediators [2]. Their role is dual-faceted: they attempt to limit infection spread and repair tissue, but persistent activation can disrupt the blood-brain barrier (BBB) integrity and exacerbate edema. Astrocytic dysfunction impairs glutamate uptake, leading to excitotoxic neuronal death, and alters metabolic support to neurons, worsening the inflammatory milieu. Oligodendrocytes, primarily responsible for myelination, are also affected. Inflammatory cytokines and oxidative stress can induce oligodendrocyte injury or apoptosis, contributing to demyelination and long-term neurological deficits commonly seen in survivors of severe meningitis and encephalitis [3]. Moreover, the sustained inflammatory environment induces a feed-forward loop where glial cells perpetuate inflammation through continuous release of cytokines, chemokines, and reactive oxygen and nitrogen species. This chronic glial activation not only damages neurons but also impairs neurogenesis and synaptic plasticity, underlying cognitive and behavioral impairments post-infection. In viral encephalitis, such as that caused by herpes simplex virus, glial cells may also present viral antigens, engaging adaptive immune responses that, while targeting the virus, may inadvertently damage host tissue. Overall, while glial activation is initially protective, excessive or prolonged activation during brain inflammation in meningitis and encephalitis leads to significant glial dysfunction, contributing to neural damage, impaired repair processes, and lasting neurological sequelae [4].

Inflammation within the brain, often triggered by injury, infection, or chronic neuroinflammatory conditions, can profoundly alter the normal processes governing glial cell development. Gliogenesis, the process by which glial cells essential for supporting neurons and maintaining brain homeostasis, are generated from neural progenitors, is tightly regulated by a network of signaling pathways and transcription factors. Among these, key regulators such as FGFR3, JAK-STAT, STAT3, S100, Hey1, HES1, DTX, IL-6, NF-κB, Neuregulin-1, MAPK-MEK, E2F/TCFL2, and NFIX/Ephrins/Netrins play crucial roles in maintaining the delicate balance between glial cell proliferation and differentiation. Dysregulation of these pathways due to brain inflammation can disrupt the normal development of glial cells, leading to an overproduction of proliferating progenitor cells and preventing their appropriate differentiation into functional glial subtypes [5]. This imbalance can have severe consequences, including an increased risk of neurodevelopmental disorders and brain tumors, particularly gliomas, which arise from glial cells. Glioma formation is strongly associated with changes in the proliferative capacity of glial progenitors and the disruption of differentiation pathways, both of which may be exacerbated by chronic brain inflammation. Understanding how brain inflammation affects these gliogenic regulators is vital to comprehending the underlying mechanisms that contribute to tumorigenesis [6]. Moreover, this research could offer new insights into potential therapeutic strategies aimed at controlling gliogenesis and mitigating the risk of brain tumors in individuals exposed to prolonged neuroinflammatory conditions. Therefore, exploring the complex interplay between brain inflammation and gliogenic regulators is essential for advancing our understanding of glial cell biology and its role in neurooncology [7, 8].

### Clinical Significance of This Study

This study holds critical clinical significance as it focuses on the long-term neurological consequences of brain inflammation, particularly in commonly encountered clinical scenarios such as meningitis and encephalitis. While the acute management of these conditions often focuses on infection control and immediate neuroprotection, this research shifts attention toward the latent risks driven by glial dysregulation. By focusing on proinflammatory cytokines such as IL-6 and TNF-α alter key gliogenic signaling pathways (e.g., JAK-STAT, FGFR3, STAT3), the study points to mechanistic links between early glial cell fate disruption and increased susceptibility to glial-derived brain tumors. These insights are especially relevant for pediatric and critical care practitioners, as young brains undergoing inflammation during developmental windows may be more vulnerable to enduring changes in glial proliferation and differentiation, potentially seeding the path toward glioma formation years later.

Furthermore, the study highlights the necessity for a paradigm shift in post-inflammatory CNS care. Glial inflammation has also been linked to many psychiatric illnesses, this study may contribute towards their better understanding as well because of the significant focus on neuro-inflammation.

Current clinical practices rarely include longitudinal surveillance of glial health following severe CNS inflammation, but the chronic activation and maladaptive responses of astrocytes, oligodendrocytes, and microglia may play a pre-neoplastic role. Understanding these processes of inflammation-induced gliogenic imbalance offers a foundation for developing early diagnostic biomarkers and therapeutic targets that can interrupt the inflammatory-to-oncogenic cascade. Thus, this study provides a compelling rationale for integrating long-term neuro-oncological risk assessment into the clinical follow-up of patients with a history of neuroinflammatory disease, ultimately aiming to improve early detection and prevention strategies in neuro-oncology.

## Methods

### Aim of the Study

With focus on improving clinical outcomes for patients, this study aims to investigate how inflammatory processes in the brain, particularly those mediated by proinflammatory signaling, disrupt key gliogenic pathways including FGFR3, JAK-STAT, STAT3, S100, Hey1, HES1, DTX, Neuregulin-1, MAPK/MEK, E2F/TCFL2, NFIX/Ephrins/Netrins thereby leading to glial cell fate dysregulation and predisposing neural tissues to tumorigenic transformation. The study focuses on how these disruptions in gliogenic regulators contribute to altered gliogenesis and increased brain tumor risk.

### Research Question

How does brain inflammation disrupt gliogenic pathways such as FGFR3, JAK-STAT, STAT3, S100, Hey1, HES1, DTX, IL-6, NF-κB, Neuregulin-1, MAPK/MEK, E2F/TCFL2, and NFIX/Ephrins/Netrins, and how do these disruptions alter glial cell fate and contribute to increased brain tumor risk? And how can this understanding contribute to the early tumor risk detection and improved patient outcomes?

### Search Focus

A comprehensive literature search was conducted using the PUBMED database, MEDLINE database, and Google Scholar, as well as open access and subscription-based journals. There were no date restrictions for published articles. The search strategy targeted:

- Neuroinflammation-Mediated Dysregulation in gliogenic regulators: FGFR3, JAK-STAT, STAT3, S100, Hey1, HES1, DTX, IL-6, NF-κB, Neuregulin-1, MAPK/MEK, E2F/TCFL2, and NFIX/Ephrins/Netrins
- Neuroinflammatory consequences for glial cell fate
- Neuroinflammation-driven reprogramming and its connection and its connection to long-term brain tumor risk

Screening of the literature was also done on this same basis and related data was extracted. Literature search began in December 2020 and ended in August 2024. An in-depth investigation was conducted during this duration based on the parameters of the study as defined above. During revision, further literature was searched and referenced until May 2025. The literature search and all sections of the manuscript were checked multiple times during the months of revision (September 2024 – May 2025) to maintain the highest accuracy possible. This comprehensive approach ensured that the selected studies provided valuable insights with focus on the objectives of the study. This study adheres to relevant PRISMA guidelines (Preferred Reporting Items for Systematic Reviews and Meta-Analyses).

### Search Queries/Keywords

1. **Inflammation and Gliogenesis Disruption:**

- “IL-6 and gliogenesis in brain inflammation”
- “NF-κB regulation of glial lineage genes”
- “Neuroinflammation and glial differentiation pathways”
- “Inflammatory signaling and STAT3 in gliogenesis”
- “Brain inflammation and brain tumor risk”
2. Gliogenic Pathways and Glial Fate Disruption:

- “FGFR3 and glial cell fate”
- “STAT3 and JAK-STAT in neural glial differentiation”
- “Notch-HES1/HEY1 axis in glial cell specification”
- “MAPK/MEK signaling in reactive gliogenesis and glioma”
- “E2F and TCFL2 transcriptional control in glial lineage regulation”
- “NFIX, Ephrins, and Netrins in glial development and tumorigenesis”
3. Tumorigenic Risk and Fate Disturbances:

- “Glial dysregulation and glioma initiation”
- “Inflammation-induced transcriptional reprogramming in the brain”
- “Disrupted gliogenesis and neural tumor predisposition”

Boolean operators (AND, OR) and field-specific filters were used to narrow down mechanistic and experimental studies linking inflammation, gliogenesis, and oncogenesis.

### Objectives of the Search

- To look into how the inflammatory mediators impact glial lineage determination and maintenance
- To trace how neuroinflammation induced disruptions in gliogenic regulators alter brain tissue homeostasis
- To investigate the inflammatory-to-oncogenic trajectory in glial cells and resultant long-term brain tumor risk.

### Screening and Eligibility Criteria

Initial Screening:

- Titles and abstracts were assessed for relevance to gliogenesis, inflammation, and brain tumor risk
- Preference was given to studies focused on disruptions in glial regulatory networks under inflammatory stress

### Full-Text Evaluation

- Focus was on studies that described direct or indirect effects of neuroinflammation on glial differentiation, reprogramming, or transformation, and associated brain tumor risk.

### Data Extraction Criteria

- Role of proinflammatory signaling in glial fate determination
- Functional consequences of dysregulated gliogenic regulators
- Evidence of gliogenic disruption predisposing to tumor formation

### Inclusion and Exclusion Criteria

Inclusion Criteria:

- Studies focused on neuroinflammation and gliogenesis
- Research focusing on gliogenic regulators (FGFR3, JAK-STAT, STAT3, S100, Hey1, HES1, DTX, IL-6, NF-κB, Neuregulin-1, MAPK/MEK, E2F/TCFL2, NFIX/Ephrins/Netrins) under inflammatory conditions
- Studies linking glial dysregulation to brain tumors Exclusion Criteria:
- Studies not involving glial cell fate or inflammation-driven mechanisms
- Research unrelated to molecular or cellular changes in glial regulators
- Articles lacking insights into the dysregulation of gliogenic regulators
- Articles that did not conform to the study focus.
- Insufficient methodological rigor.

### Rationale for Screening and Inclusion

The gliogenic regulators selected are core to gliogenic programming and are known to be sensitive to inflammatory signals. Among them, STAT3, FGFR3, and NFIX have dual roles in normal gliogenesis and glioma susceptibility. Notch pathway members (HES1, HEY1), along with IL-6/NF-κB, mediate inflammation-induced reactive gliosis, which may serve as a precursor state to tumorigenic conversion. The integration of signaling (MAPK/MEK, JAK-STAT, E2F) and cell fate determination factors under inflammatory stress is essential for understanding the trajectory from developmental dysregulation to associated brain tumor risk.

**FGFR3** – Included for its pivotal role in astrocyte differentiation and its inflammatory dysregulation linked to glioma progression.

**JAK-STAT** – Selected due to its central role in cytokine signaling during inflammation and its control over gliogenesis and glial cell proliferation.

**STAT3** – Included for its dual function in glial lineage specification and inflammatory response amplification that may predispose to oncogenic transformation.

**S100** – Chosen due to its calcium-binding role in glial maturation and inflammatory regulation, often elevated in glial pathologies.

**Hey1** – Included for its role as a Notch target in gliogenesis, sensitive to inflammatory cues that shift progenitor fate.

**HES1** – Selected as a critical Notch effector in glial fate decisions, dysregulated under chronic neuroinflammation. **DTX** – Included for its involvement in modulating Notch signaling, influencing glial cell outcomes under inflammatory conditions.

**IL-6** – Selected due to its inflammatory cytokine role in promoting reactive gliosis and glial proliferation, contributing to tumor-prone environments.

**NF-κB** – Included for its key role in mediating inflammatory gene expression and affecting glial cell survival, proliferation, and tumorigenesis.

**Neuregulin-1** – Selected due to its influence on glial differentiation and response to inflammatory signaling disruptions.

**MAPK/MEK** – Included for their integration of mitogenic and inflammatory signals affecting glial cell cycle progression and fate.

**E2F/TCFL2** – Included for their regulation of glial progenitor proliferation and vulnerability to inflammation-induced transcriptional dysregulation.

**NFIX/Ephrins/Netrins** – Included for their roles in spatial guidance and gliogenic patterning, susceptible to inflammatory disruption impacting glial positioning and transformation.

PRISMA Flow Diagram is Fig 1.

**Fig 1.**
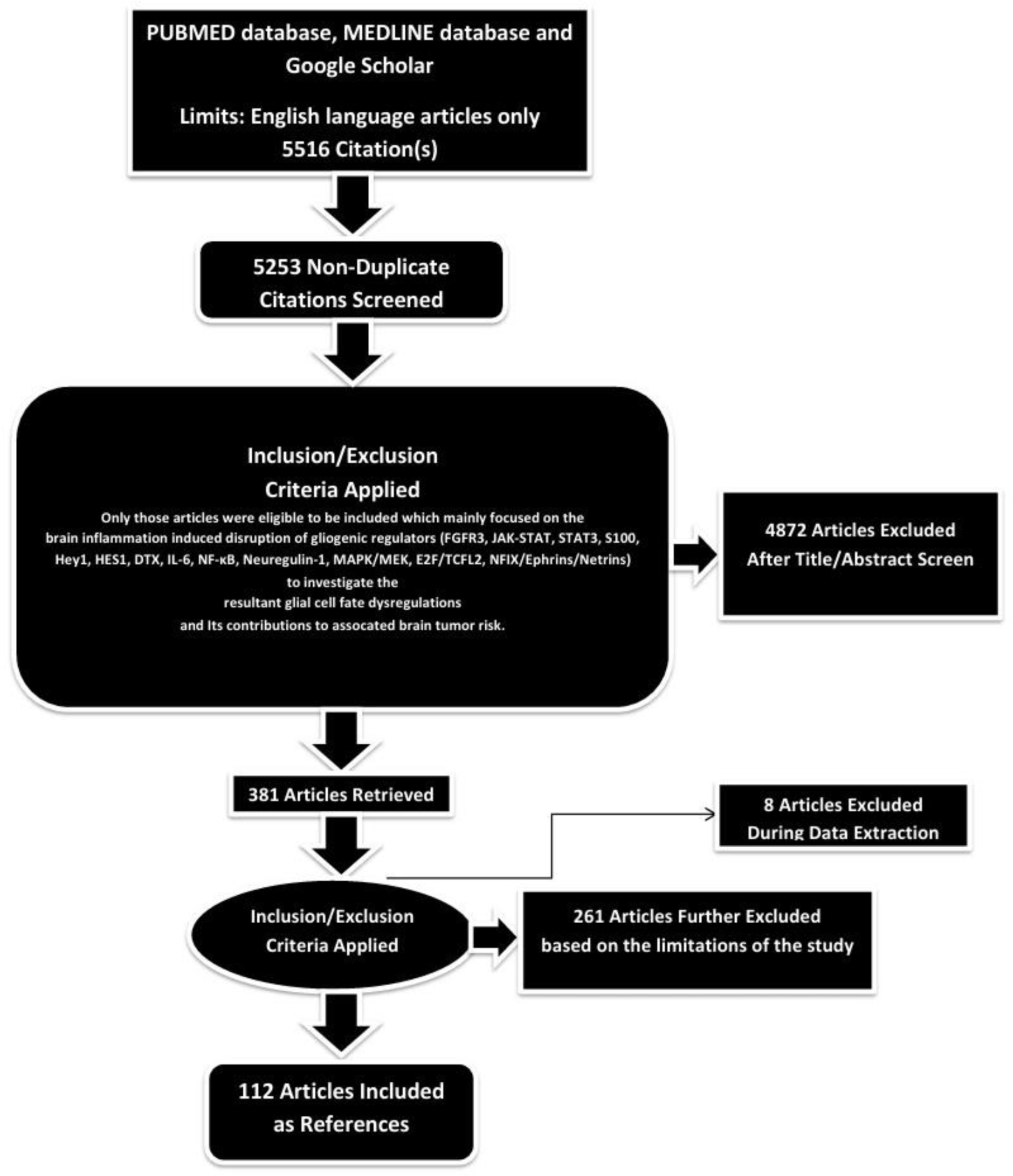
PRISMA FLOW DIAGRAM. This figure represents graphically the flow of citations in the study.

### Assessment of Article Quality and Potential Biases

Ensuring the quality and minimizing potential biases of the selected articles were crucial aspects to guarantee the rigor and reliability of the research findings.

### Quality Assessment

The initial step in quality assessment involved evaluating the methodological rigor of the selected articles. This included a thorough examination of the study design, data collection methods, and analyses conducted. The significance of the study’s findings was weighed based on the quality of the evidence presented. Articles demonstrating sound methodology such as well-designed studies, controlled variables, and scientifically robust data were considered of higher quality. Methodological rigor served as a significant indicator of quality.

### Potential Biases Assessment

- **Publication Bias:** To address the potential for publication bias, a comprehensive search strategy was adopted to include a balanced representation of both positive and negative results, incorporating a wide range of published articles from databases like Google Scholar.
- **Selection Bias:** Predefined and transparent inclusion criteria were applied to minimize subjectivity in the selection process.

Articles were chosen based on their relevance to the study’s objectives, adhering strictly to these criteria. This approach reduced the risk of subjectivity and ensured that the selection process was objective and consistent.

- **Reporting Bias:** To mitigate reporting bias, articles were checked for inconsistencies or missing data. Multiple detailed reviews of the methodologies and results were conducted for all selected articles to identify and address any reporting bias.

By including high-quality studies and thoroughly assessing potential biases, this study aimed to provide a robust foundation for the results and conclusions presented.

### Language and Publication Restrictions

We restricted our selection to publications in the English language. There were no limitations imposed on the date of publication. Unpublished studies were not included in our analysis.

By investigating brain inflammation induced disruption of gliogenic regulators and resultant glial cell fate dysregulations, this study provides significant insights into the mechanisms driving associated brain tumor risk and associated clinical implications.

## Results

A total of 5516 articles were identified using database searching, and 5253 were recorded after duplicates removal. 4872 were excluded after screening of title/abstract, 261 were finally excluded, and 8 articles were excluded during data extraction.

Finally, 112 articles were included as references.

### Clinical Relevance and Importance Towards Improving Patient Outcomes

This study carries substantial clinical relevance as it focuses on how neuroinflammation-induced dysregulation of gliogenic regulators such as FGFR3, STAT3, IL-6, and NF-κB, can impair normal glial cell differentiation and lead to conditions that favor tumorigenesis. In clinical settings, patients suffering from meningitis, encephalitis, or chronic neuroinflammatory disorders often receive care focused on acute infection control and symptomatic relief. However, this study emphasizes that inflammatory insults to the central nervous system may have lasting impacts on glial biology, altering cell fate pathways and increasing the long-term risk of glial-derived brain tumors such as gliomas. These findings are particularly important for vulnerable populations, including pediatric and immunocompromised patients, who may face a higher likelihood of incomplete glial recovery and unrecognized tumor susceptibility following severe CNS inflammation.

Translating this knowledge into clinical practice has the potential to significantly improve patient outcomes. By identifying the molecular markers and signaling networks that are disrupted during inflammation such as the JAK-STAT and Notch-HES1/HEY1 axes, this study also has the potential to possibly contribute to the future interventions aimed at preserving glial homeostasis. Therapeutic targeting of these dysregulated pathways during or after neuroinflammatory episodes could prevent maladaptive glial proliferation and reduce tumor risk. Moreover, this study supports the implementation of long-term surveillance strategies for patients with a history of neuroinflammation, incorporating molecular risk profiling and neuroimaging to monitor glial integrity. Ultimately, these preventative and personalized approaches could shift the current model of care from reactive treatment to proactive risk reduction, thereby improving neurological outcomes and reducing brain tumor incidence in inflammation-exposed populations.

### Investigating Brain Inflammation-Induced Dysregulation of Gliogenic Signaling and Associated Brain Tumor Risk

#### 1. FGFR3

##### Neuroinflammation-Mediated Dysregulation in Glial Cells

Fibroblast growth factor receptor 3 (FGFR3) plays a crucial role in gliogenesis, particularly in the specification and maturation of astrocytes and oligodendrocyte progenitor cells during brain development. Under normal physiological conditions, FGFR3 activation by its ligands, such as FGF1 and FGF2, orchestrates controlled signaling through downstream pathways like MAPK/ERK and PI3K/AKT, promoting glial differentiation, maintaining progenitor cell pools, and regulating proliferation and survival [9]. However, during brain inflammation, such as that occurring in meningitis and encephalitis, the tightly regulated expression and activation of FGFR3 can become severely dysregulated. Pro-inflammatory cytokines like IL-1β, TNF-α, and IL-6, which are abundantly produced during neuroinflammation, can directly or indirectly interfere with FGFR3 signaling. These cytokines can alter the transcriptional regulation of the FGFR3 gene, either suppressing its expression through inflammatory transcription factors like NF-κB or modifying epigenetic marks on the FGFR3 promoter, leading to aberrant gene silencing or inappropriate activation [10].

Inflammation-induced oxidative stress and the release of reactive oxygen species (ROS) further contribute to FGFR3 pathway dysregulation. Oxidative modifications of signaling intermediates or FGFR3 itself can impair receptor function, while prolonged inflammation can shift the balance of ligand availability, favoring conditions that either overstimulate or insufficiently activate FGFR3. For instance, elevated FGF2 levels in an inflamed environment may cause excessive FGFR3 activation, leading to abnormal glial proliferation and astrogliosis rather than organized gliogenesis [11]. Conversely, inflammation-mediated damage to the extracellular matrix and impaired ligand production can attenuate FGFR3 signaling, resulting in reduced astrocyte and oligodendrocyte formation at critical stages of brain repair. Moreover, chronic inflammatory signals can cause downstream desensitization of FGFR3 signaling pathways, with inhibitory proteins like SOCS (suppressors of cytokine signaling) being upregulated and interfering with normal receptor-mediated responses. MicroRNAs induced during inflammation may also target FGFR3 transcripts, reducing their stability or translation efficiency and further dampening the receptor’s function. Additionally, inflammatory environments may favor alternative splicing events of FGFR3 pre-mRNA, producing isoforms with altered signaling capacities that do not support normal gliogenic outcomes. Collectively, these dysregulations disturb the delicate orchestration of gliogenesis, leading to defective glial cell maturation, improper responses to injury, and contributing to the long-term neuropathology observed after severe brain inflammation. The cumulative effect is a disruption in the glial supportive network essential for neuronal function and brain homeostasis, setting the stage for persistent neurological deficits and impaired brain recovery [12].

##### Consequences for Glial Cell Fate and Long-Term Brain Tumor Risk

Brain inflammation, particularly during critical developmental periods or following severe infections like meningitis and encephalitis, can profoundly disrupt FGFR3 signaling, thereby altering glial cell fate decisions in ways that may increase the risk of brain tumor formation later in life [13]. Under normal conditions, FGFR3 tightly regulates the proliferation, differentiation, and maturation of glial progenitor cells, guiding their commitment toward functional astrocytes and oligodendrocytes. However, inflammatory insults can dysregulate FGFR3 at multiple levels, leading to aberrant signaling that disrupts the balance between proliferation and differentiation. Elevated levels of pro-inflammatory cytokines and oxidative stress can interfere with FGFR3 activation and its downstream signaling cascades, such as the MAPK/ERK and PI3K/AKT pathways, both of which are critical for maintaining proper glial lineage specification. As a result, glial progenitor cells may either fail to fully differentiate or remain in an undifferentiated, proliferative state, creating a pool of cells that are more susceptible to malignant transformation [14]. The sustained inflammatory environment can also promote genetic and epigenetic changes in glial progenitor cells, including mutations or aberrant methylation patterns affecting FGFR3 itself or its key regulatory networks. Such alterations may further enhance uncontrolled proliferation and impair normal cell cycle checkpoints. In some cases, chronic inflammation may lead to gain-of-function mutations in FGFR3, resulting in constitutive activation of the receptor independent of ligand binding. This persistent FGFR3 signaling can drive unchecked cell division and inhibit normal differentiation, two hallmarks of oncogenic processes. In addition, the presence of inflammatory mediators such as IL-6 and TNF-α can synergize with dysregulated FGFR3 signaling to activate oncogenic transcription factors like STAT3, further promoting a pro-tumorigenic environment. Another important consequence of inflammation-driven FGFR3 dysregulation is the alteration of the local microenvironment, including breakdown of the blood-brain barrier, chronic astrocyte activation, and persistent recruitment of immune cells, all of which contribute to a milieu that supports neoplastic transformation. Oligodendrocyte progenitor cells, which naturally express high levels of FGFR3 during development, are particularly vulnerable [15]. If their normal maturation pathways are disrupted, they may persist as an expanded, aberrant progenitor pool capable of giving rise to tumors such as gliomas or, more specifically, oligodendrogliomas. Additionally, since FGFR3 alterations have been implicated in the formation of certain low-grade gliomas and mixed neuronal-glial tumors, early-life inflammatory disruptions to this pathway could set the molecular groundwork for tumorigenesis that manifests clinically years or decades later. Overall, brain inflammation’s capacity to interfere with FGFR3 signaling not only disrupts the normal developmental trajectories of glial cells but also creates a cellular and molecular landscape ripe for oncogenic transformation [16].

##### Possible Clinical Implications in Patients: Toward Improved Diagnosis, Surveillance, and Therapeutics

The dysregulation of FGFR3 signaling during neuroinflammatory episodes presents a compelling opportunity to enhance diagnostic precision and long-term surveillance in patients recovering from CNS infections such as meningitis and encephalitis. Given FGFR3’s central role in glial lineage specification and its vulnerability to inflammatory cytokines and oxidative stress, monitoring FGFR3 activity or expression patterns could serve as an early biomarker for glial dysregulation in post-infection states. Diagnostic panels incorporating FGFR3-related molecular alterations such as aberrant phosphorylation, alternative splicing isoforms, or inflammation-induced microRNA targeting could aid clinicians in stratifying patients by their risk for delayed glial recovery or progression toward a pre-neoplastic state.

Additionally, imaging strategies targeting glial cell metabolism and density in FGFR3-rich brain regions may provide non-invasive tools for early detection of glial pathology before structural abnormalities appear. From a therapeutic standpoint, targeting FGFR3 dysregulation during or after neuroinflammatory events may hold promise in preserving healthy gliogenesis and preventing oncogenic transformation. Anti-inflammatory interventions aimed at limiting IL-6, TNF-α, or NF-κB activity could help maintain the fidelity of FGFR3 signaling pathways. More specifically, FGFR3-modulating agents such as ligand analogs or selective kinase inhibitors, could be explored to restore balanced proliferation and differentiation in glial progenitors. Furthermore, integrating FGFR3-related molecular profiling into follow-up protocols for pediatric or immunocompromised patients who experience CNS inflammation could enable early therapeutic intervention before glioma initiation. By addressing the chronic impact of inflammation on FGFR3, clinicians may be able to reduce the incidence of glial-derived tumors and improve long-term neurological outcomes in high-risk patient populations.

#### 2. JAK-STAT Pathway

##### Neuroinflammation-Mediated Dysregulation in Glial Cells

The JAK-STAT signaling pathway plays a vital role in gliogenesis, particularly in promoting the differentiation of neural stem cells into astrocytes during brain development. Under physiological conditions, cytokines such as leukemia inhibitory factor (LIF) and ciliary neurotrophic factor (CNTF) activate the Janus kinases (JAKs), which then phosphorylate and activate signal transducer and activator of transcription (STAT) proteins, primarily STAT3.

Activated STAT3 translocates into the nucleus and drives the transcription of genes necessary for astrocyte fate commitment and maturation [17]. However, during brain inflammation, as seen in meningitis, encephalitis, or other neuroinflammatory conditions, the regulation of the JAK-STAT pathway can become significantly disrupted. The excessive presence of pro-inflammatory cytokines such as IL-6, interferons, and TNF-α massively overstimulates the pathway, leading to hyperactivation of STAT3 and other STAT family members. This persistent activation skews the delicate balance between neurogenesis and gliogenesis, resulting in exaggerated astrocyte formation at the expense of neurons and oligodendrocytes, thereby altering normal brain development and repair processes. Beyond hyperactivation, brain inflammation can also induce negative feedback mechanisms that paradoxically suppress JAK-STAT signaling over time [18]. For instance, the expression of suppressor of cytokine signaling (SOCS) proteins, particularly SOCS3, is upregulated in response to chronic JAK-STAT activation. SOCS3 binds to JAKs and prevents further phosphorylation of STATs, effectively shutting down the signaling cascade. In an inflammatory environment, the overexpression of SOCS3 can prematurely inhibit JAK-STAT signaling, leading to an inadequate astrocytic response or defective glial maturation during critical periods of brain development or repair. Moreover, oxidative stress generated during inflammation can modify JAK and STAT proteins post-translationally, impairing their function or stability and further contributing to pathway dysregulation. Epigenetic changes, such as DNA methylation or histone modifications at the promoters of JAKs, STATs, or their target genes, may also arise in the inflamed brain, leading to long-term silencing or inappropriate activation of key components necessary for normal gliogenesis. Inflammatory microenvironments can additionally alter the cellular responsiveness to JAK-STAT signaling by modifying the expression or localization of cytokine receptors on glial progenitor cells. Downregulation or internalization of receptors like gp130, critical for LIF and CNTF signaling, diminishes the ability of cells to respond appropriately to differentiation cues. In parallel, microRNAs induced by inflammation, such as miR-21 and miR-155, may directly target JAK-STAT pathway transcripts, reducing their levels and further disrupting normal signaling dynamics [19]. As a result, glial progenitor cells may fail to properly differentiate, or they may adopt abnormal reactive phenotypes that are not conducive to healthy brain structure and function. Thus, brain inflammation exerts a profound and multifaceted impact on JAK-STAT pathway regulation, disrupting the careful orchestration of gliogenesis and contributing to both immediate developmental disturbances and long-term vulnerabilities in the central nervous system[20].

##### Consequences for Glial Cell Fate and Long-Term Brain Tumor Risk

Brain inflammation can severely impact the JAK-STAT signaling pathway, leading to disruptions in the normal cell fate determination of glial cells and setting the stage for an increased risk of brain tumor development later in life. In the healthy developing brain, the JAK-STAT pathway, particularly through STAT3 activation, is essential for guiding neural progenitor cells toward an astrocytic lineage in a tightly regulated and temporally controlled manner [21]. However, during episodes of inflammation such as meningitis, encephalitis, or chronic neuroinflammatory conditions, the inflammatory milieu, rich in cytokines like IL-6, IL-1β, and interferons, induces hyperactivation of the JAK-STAT pathway. This exaggerated activation can force an excessive commitment of progenitor cells toward astrocyte-like fates, resulting in an abnormal expansion of reactive astrocyte populations. At the same time, this hyperactivation may inhibit oligodendrocyte and neuronal differentiation pathways, leading to an imbalance in the cellular composition of the brain and a population of glial cells that retain proliferative and plastic characteristics beyond their normal developmental windows [22]. The persistence of an abnormally expanded glial progenitor pool under the influence of chronic inflammatory stimuli increases the risk of oncogenic transformation. Hyperactivated STAT3 signaling is well-recognized not only for its role in astrocytic differentiation but also for its ability to promote cell proliferation, survival, and immune evasion, hallmarks of cancer cells. Prolonged inflammation-induced activation of STAT3 can therefore drive glial cells into a pre-neoplastic state, characterized by unchecked proliferation and resistance to normal regulatory mechanisms. In addition, chronic inflammation leads to the upregulation of negative regulators like SOCS3, which initially act to curb excessive JAK-STAT signaling but may also create periods of instability in the pathway’s regulation. This instability can allow for the accumulation of genetic mutations, epigenetic alterations, and aberrant activation of oncogenic programs in glial progenitor cells. Furthermore, inflammatory environments promote oxidative stress and DNA damage, compounding the risk that glial cells with dysregulated JAK-STAT signaling will acquire additional mutations necessary for full malignant transformation. In particular, brain regions rich in progenitor populations, such as the subventricular zone, become susceptible to giving rise to tumor-initiating cells. Glioblastomas and astrocytomas, two of the most aggressive forms of brain tumors, have been associated with persistent activation of the JAK-STAT3 axis, supporting the idea that early-life inflammatory disruption of this pathway can lay the groundwork for tumorigenesis that may manifest years or decades later [23]. Additionally, microRNAs induced during inflammation, such as miR-21, can further enhance STAT3 signaling while suppressing tumor suppressor pathways, providing a molecular bridge between inflammation, aberrant glial fate, and tumor initiation. Overall, the impact of brain inflammation on the JAK-STAT pathway creates a cascade of events that derails the normal fate decisions of glial cells, fosters an environment of chronic proliferative signaling, and introduces genetic and epigenetic instability. Together, these factors significantly elevate the lifetime risk of developing glial-derived brain tumors, highlighting the importance of protecting the developing and injured brain from inflammatory insults to preserve long-term neurological health [24].

##### Possible Clinical Implications in Patients: Toward Improved Diagnosis, Surveillance, and Therapeutics

Pathway Dysregulation of the JAK-STAT pathway in the context of brain inflammation presents clinically actionable insights for improving diagnosis and long-term care of patients who experience CNS infections or neuroinflammatory episodes. Since hyperactivation of STAT3 during inflammation is known to drive excessive astrocytic differentiation while impairing oligodendrocyte and neuronal development, early detection of pathway imbalances, using phosphorylated STAT3 levels, cytokine profiles (e.g., IL-6, IFN-γ), or SOCS3 expression, could serve as valuable biomarkers for identifying patients at risk of abnormal glial fate decisions. In pediatric patients, where brain development is especially sensitive to inflammatory disruption, these biomarkers could support early diagnosis of gliogenic imbalance and guide clinicians toward more personalized follow-up. Molecular surveillance of JAK-STAT signaling status in patients recovering from meningitis, encephalitis, or chronic neuroinflammatory diseases could also help identify those at higher risk of developing glial-related tumors later in life, particularly in populations with genetic predispositions or in regions of the brain rich in progenitor pools. Therapeutically, modulating the JAK-STAT axis offers a promising avenue to protect glial integrity and reduce oncogenic risk following inflammation.

Pharmacologic inhibitors of STAT3, or regulators that enhance SOCS3 precision without inducing full pathway collapse, could help rebalance the overstimulated gliogenic environment in inflamed brains. Anti-cytokine therapies, such as IL-6 blockers (e.g., tocilizumab), may serve as adjuncts in acute CNS inflammation to minimize exaggerated JAK-STAT activation. Furthermore, epigenetic modulators or miRNA-targeting therapeutics that restore proper STAT3 expression and activity could be explored to reverse long-term glial dysregulation in at-risk individuals. This study also highlights the clinical need for routine cognitive and neuroimaging surveillance in patients with a history of neuroinflammation, particularly those with persistent gliosis or abnormal glial activity. By addressing the molecular aftermath of JAK-STAT pathway dysregulation, clinicians may not only preserve brain development and repair capacity but also reduce the lifetime risk of astrocytoma or glioblastoma formation, ultimately improving long-term neurological outcomes.

#### 3. STAT3

##### Neuroinflammation-Mediated Dysregulation in Glial Cells

STAT3 plays a central role in gliogenesis, particularly in promoting the differentiation of neural stem and progenitor cells into astrocytes during brain development. Its activation is tightly regulated under normal physiological conditions by extracellular cues such as cytokines, including leukemia inhibitory factor (LIF) and ciliary neurotrophic factor (CNTF), which engage receptors that activate associated Janus kinases (JAKs), leading to the phosphorylation and activation of STAT3 [25]. Phosphorylated STAT3 dimerizes and translocates to the nucleus where it regulates the transcription of genes critical for astrocyte maturation and function. However, brain inflammation disrupts this carefully regulated signaling process in multiple ways. In the inflamed brain, high levels of cytokines such as IL-6, IL-1β, and TNF-α result in sustained and excessive activation of STAT3. This hyperactivation can disturb the normal timing and extent of astrocyte differentiation, prematurely pushing progenitor cells towards astrocytic fates at the expense of neuronal and oligodendrocyte lineages. In addition to hyperactivation, brain inflammation can also provoke negative regulatory feedback loops that inhibit STAT3 signaling over time [26]. One key mechanism is the upregulation of suppressor of cytokine signaling 3 (SOCS3), which is itself a transcriptional target of STAT3. SOCS3 binds to JAKs and associated cytokine receptors, blocking further STAT3 activation. During chronic inflammation, the overexpression of SOCS3 can suppress STAT3 activity to suboptimal levels, impairing the normal differentiation of astrocytes and potentially leaving progenitor cells in a dysfunctional or undifferentiated state. Moreover, oxidative stress generated during inflammation can induce modifications of STAT3, such as nitration or oxidation, altering its ability to be phosphorylated, to dimerize properly, or to bind DNA effectively. Such post-translational modifications can impair STAT3’s normal gene regulatory functions and lead to either loss of astrocyte-supportive gene expression or aberrant expression of genes that promote reactive, maladaptive glial phenotypes. Inflammation can also reshape the epigenetic landscape of STAT3 and its target genes [27]. Pro-inflammatory signals can induce changes in DNA methylation and histone modifications around STAT3-binding sites, leading to sustained transcriptional repression or inappropriate activation of specific genes involved in gliogenesis. Inflammatory conditions may also alter the expression of non-coding RNAs, such as microRNAs, that target STAT3 mRNA for degradation or inhibit its translation. For example, inflammation-induced microRNAs like miR-124 and miR-21 have been implicated in modulating STAT3 levels and activity, adding another layer of complexity to how inflammation disturbs STAT3-dependent glial development. Moreover, inflammatory environments can alter the expression of cell surface receptors that mediate STAT3 activation, such as gp130 and CNTF receptors, diminishing the ability of neural progenitor cells to properly respond to developmental cues. This impaired receptor signaling, combined with aberrant intracellular regulation of STAT3, can desynchronize the temporal progression of gliogenesis, potentially leading to the expansion of abnormal, reactive, or proliferative glial populations. Collectively, brain inflammation exerts a multifaceted dysregulation of STAT3 gene expression and signaling dynamics, disrupting the finely tuned developmental processes necessary for generating healthy glial populations and maintaining central nervous system homeostasis [28].

##### Consequences for Glial Cell Fate and Long-Term Brain Tumor Risk

Brain inflammation exerts profound effects on the STAT3 signaling pathway, leading to disruptions in glial cell fate determination that can ultimately increase the risk of brain tumor development later in life. Under normal developmental conditions, STAT3 activation is critical for promoting the differentiation of neural stem cells into astrocytes, a process tightly regulated to ensure balanced production of glial and neuronal populations. However, during episodes of brain inflammation, an excess of pro-inflammatory cytokines such as IL-6, IL-1β, and TNF-α leads to sustained and often exaggerated activation of STAT3 [29]. This hyperactivation accelerates and biases the differentiation of progenitor cells toward astrocytic fates at the expense of neuronal and oligodendroglial lineages, disrupting the normal cell fate balance. Furthermore, the persistent activation of STAT3 under inflammatory conditions can push astrocyte progenitors into a reactive state characterized by hypertrophy, upregulation of inflammatory genes, and loss of normal supportive functions, all of which prime the cells for pathological remodeling rather than stable maturation. Chronic inflammatory conditions also create an environment where glial progenitor cells are exposed to continuous proliferative and survival signals mediated through STAT3 [30]. While these signals are protective in acute injury settings, their persistent activation can promote the accumulation of undifferentiated or partially differentiated astrocyte-like cells with heightened proliferative capacity and genomic instability. Moreover, chronic STAT3 signaling enhances the expression of genes associated with oncogenic transformation, such as Cyclin D1 and Bcl-XL, while simultaneously repressing pro-apoptotic signals. This dual effect creates a cellular environment conducive to the survival and expansion of cells harboring oncogenic mutations, thereby laying the groundwork for future tumor formation. Inflammatory-induced oxidative stress further exacerbates the problem by inducing DNA damage in proliferating glial progenitors. In the presence of dysregulated STAT3 activity, mechanisms that normally halt the proliferation of damaged cells become ineffective, allowing the clonal expansion of mutated cells.

Additionally, persistent inflammatory signaling can alter the expression of critical negative regulators of STAT3, such as SOCS3, leading to fluctuating and unstable STAT3 activity that destabilizes glial cell identity and promotes the emergence of aberrant, tumorigenic glial populations. Epigenetic modifications induced by inflammation, such as changes in DNA methylation or histone acetylation at STAT3 target gene loci, can also lock glial cells into a dysregulated, pro-oncogenic state, making it increasingly difficult for them to revert to normal, regulated differentiation programs [31]. This cascade of events significantly elevates the risk of glioma development later in life.

In fact, high levels of activated STAT3 are commonly found in glioblastoma multiforme, a highly aggressive brain tumor, where STAT3 functions as a key driver of tumor initiation, progression, and resistance to therapy. Thus, inflammation-induced dysregulation of STAT3 not only alters the immediate trajectory of glial cell fate decisions during development and repair but also seeds a population of cells with the proliferative, survival, and genomic instability features necessary for tumorigenesis. The long-term consequences of early-life or chronic brain inflammation therefore include an increased risk of developing malignant glial tumors, underscoring the critical need to protect the developing brain from inflammatory insults and to understand the molecular pathways through which inflammation predisposes to oncogenic transformation [32].

##### Possible Clinical Implications in Patients: Toward Improved Diagnosis, Surveillance, and Therapeutics

Dysregulation of the STAT3 pathway during neuroinflammation has profound clinical implications for early diagnosis and risk stratification in patients recovering from CNS infections or inflammatory conditions. STAT3 is a central node in astrocytic differentiation and glial homeostasis, and its persistent hyperactivation, triggered by cytokines such as IL-6 and TNF-α, can serve as a molecular marker of maladaptive gliogenesis. Clinically, elevated phosphorylated STAT3 (pSTAT3) levels in cerebrospinal fluid or peripheral blood could be investigated as a biomarker for gliosis severity, glial imbalance, or even pre-oncogenic glial activation, particularly in pediatric patients or those with recurring episodes of encephalitis or meningitis. Furthermore, combining STAT3 activity profiling with imaging-based assessments of reactive gliosis may enhance surveillance protocols in patients with prior neuroinflammatory insults, allowing for earlier detection of persistent glial abnormalities that may precede tumorigenesis. From a therapeutic perspective, targeting dysregulated STAT3 signaling offers a promising avenue to interrupt the cascade of inflammation-driven gliomagenesis. Small molecule inhibitors of STAT3 or its upstream activators (such as JAKs or IL-6/gp130 complexes) could be explored as adjunct therapies during the acute or subacute phases of CNS inflammation to normalize glial differentiation patterns and reduce astrocytic hyperproliferation. Anti-inflammatory biologics, including IL-6 receptor antagonists or STAT3-targeting antisense oligonucleotides, may also help restore physiological regulation of glial progenitor fate. Importantly, the chronic consequences of STAT3 dysregulation such as its role in promoting proliferation, survival, and epigenetic reprogramming highlight the need for long-term molecular surveillance of high-risk patients. Monitoring STAT3-driven gene signatures (e.g., Bcl-XL, Cyclin D1) and SOCS3 levels over time could inform therapeutic responsiveness and risk of malignant transformation. Integrating these diagnostic and therapeutic strategies could dramatically improve outcomes by enabling earlier intervention, reducing glioma risk, and preserving neurodevelopmental integrity in patients recovering from neuroinflammatory diseases.

#### 4. S100

##### Neuroinflammation-Mediated Dysregulation in Glial Cells

The S100 family of proteins, particularly S100B, plays important roles in gliogenesis, primarily by influencing the maturation, proliferation, and function of astrocytes during brain development. S100B is expressed at high levels in maturing astrocytes and serves as both an intracellular regulator and an extracellular signaling molecule [33]. Under normal conditions, S100B promotes cytoskeletal organization, cell survival, and neurite extension, contributing to the formation of a functional glial network that supports neuronal development and homeostasis. However, during brain inflammation, the gene expression and functional behavior of S100 proteins can become profoundly dysregulated.

Pro-inflammatory cytokines such as IL-1β, TNF-α, and IL-6, as well as oxidative stress and reactive oxygen species generated during inflammation, can directly upregulate the transcription of S100B and other S100 proteins through the activation of transcription factors like NF-κB and AP-1. This upregulation often results in aberrantly high levels of extracellular S100B, which shifts its role from a trophic factor to a damage-associated molecular pattern (DAMP) molecule. At elevated concentrations, extracellular S100B interacts with receptors such as the receptor for advanced glycation end-products (RAGE), triggering chronic activation of inflammatory pathways and further amplifying cytokine production [34]. This feedback loop not only perpetuates neuroinflammation but also disrupts the normal developmental roles of S100B, leading to altered astrocyte maturation and function. Furthermore, persistent activation of the RAGE pathway by elevated S100B levels promotes the expression of additional pro-inflammatory genes, exacerbating gliosis and disturbing the balance between supportive astrocyte states and reactive, pathogenic phenotypes. Inflammation can also induce post-translational modifications of S100 proteins, such as oxidation, nitration, or phosphorylation, which alter their binding affinities and functional activities. These modifications can impair the intracellular roles of S100B in calcium buffering, cytoskeletal regulation, and cell proliferation control, leading to dysfunctional astrocyte behavior. Moreover, chronic inflammatory environments may lead to epigenetic changes, including histone modifications and DNA methylation changes at the S100B gene locus, which can lock glial cells into states of persistently high S100B expression even after the inflammatory insult has subsided. Such epigenetic dysregulation can disturb the long-term stability of astrocyte identity and contribute to maladaptive responses during later brain development and aging. In addition to S100B, other S100 family members such as S100A1, S100A6, and S100A10, which also have roles in gliogenesis and glial function, may similarly become dysregulated during inflammation [35]. These proteins are involved in processes like cell cycle regulation, response to stress, and interaction with cytoskeletal elements, and their dysregulation further compounds the destabilization of glial networks. Collectively, brain inflammation induces a multifaceted dysregulation of S100 gene expression and protein function, disrupting the normal progression of gliogenesis and creating an environment of glial dysfunction that can have lasting impacts on brain structure and function [36].

##### Consequences for Glial Cell Fate and Long-Term Brain Tumor Risk

Brain inflammation significantly alters the expression and function of S100 proteins, particularly S100B, leading to disruptions in glial cell fate that may predispose to brain tumor development later in life. Under normal developmental conditions, S100B acts as a key regulator of astrocyte maturation and cytoskeletal dynamics, promoting the transition of neural progenitor cells into functional, supportive astrocytes. However, in the context of brain inflammation, elevated levels of inflammatory cytokines such as IL-1β, TNF-α, and IL-6 drive the overexpression of S100B and related S100 family members. This pathological upregulation, often sustained beyond the resolution of the initial inflammatory insult, shifts the role of S100B from a developmental modulator to a pro-inflammatory mediator that exacerbates cellular stress responses and reactive gliosis [37]. As a result, the differentiation trajectory of glial progenitor cells becomes skewed, favoring the generation of hypertrophic, reactive astrocytes over healthy, functional glia. The chronic overproduction of S100B, particularly in its extracellular form, activates the receptor for advanced glycation end-products (RAGE) on glial cells and neighboring neurons, perpetuating a self-reinforcing loop of inflammation and cellular stress. Persistent RAGE signaling alters intracellular pathways governing cell cycle control, apoptosis, and differentiation, pushing glial progenitors toward aberrant proliferative states instead of terminal differentiation. Moreover, high levels of S100B disrupt the tightly regulated intracellular calcium signaling necessary for proper glial maturation, leading to cytoskeletal abnormalities and defects in cellular polarity. This misregulation creates a population of glial cells that retain a proliferative, immature phenotype, a known risk factor for tumorigenic transformation. Over time, this population of dysregulated glial progenitors becomes increasingly susceptible to accumulating genetic and epigenetic mutations, particularly in the pro-inflammatory environment maintained by persistent S100B-RAGE signaling. Oxidative stress induced by chronic inflammation further damages the DNA of proliferating glial cells, and under conditions of impaired differentiation and heightened S100B expression, damaged cells may evade normal apoptotic mechanisms [38]. This scenario promotes clonal expansion of mutated glial populations with tumorigenic potential. Additionally, the long-term epigenetic modifications triggered by inflammation, such as altered methylation patterns at the S100B locus or at genes regulating cell cycle and DNA repair, can entrench a dysregulated glial state that persists into adulthood. This epigenetic instability increases the likelihood that glial progenitor cells will undergo malignant transformation under additional environmental or genetic stresses later in life. The link between chronically elevated S100B levels and brain tumor risk is supported by observations in glioma and astrocytoma, where high S100B expression correlates with increased tumor grade and poor prognosis. Thus, brain inflammation’s impact on S100 proteins not only disrupts immediate glial cell fate decisions but also lays the molecular and cellular groundwork for tumorigenesis [39, 40]

##### Possible Clinical Implications in Patients: Toward Improved Diagnosis, Surveillance, and Therapeutics

The inflammation-mediated dysregulation of S100 proteins, especially S100B, has significant clinical implications for enhancing early diagnosis and long-term risk assessment in patients recovering from CNS inflammation such as meningitis, encephalitis, or traumatic brain injury. S100B, which under normal conditions promotes astrocyte maturation and neuroprotection, becomes a pro-inflammatory damage-associated molecule when overexpressed. Clinically, elevated extracellular S100B levels detectable in serum or cerebrospinal fluid, could serve as a non-invasive biomarker of astrocytic distress, reactive gliosis, or unresolved neuroinflammation. Such measurements, particularly when tracked longitudinally, could help clinicians identify patients who are at risk of persistent glial dysregulation and future neuro-oncological complications. Moreover, S100B-driven activation of RAGE and its downstream signaling cascade could be targeted in surveillance protocols by assessing the presence of RAGE-positive glial populations or inflammatory mediators indicative of chronic S100 signaling. This would allow for the identification of patients who may benefit from early anti-inflammatory or neuroprotective interventions aimed at restoring glial homeostasis.

Therapeutically, strategies aimed at modulating S100B expression or blocking its interaction with RAGE hold potential in reducing the chronic inflammatory feedback loop that primes glial cells for oncogenic transformation. RAGE antagonists or inhibitors of NF-κB activation may disrupt this vicious cycle and help prevent maladaptive astrocyte proliferation and gliosis. Antioxidants and agents that stabilize intracellular calcium signaling may also mitigate the cytoskeletal disorganization and proliferative shift seen in S100B-dysregulated astrocytes. Additionally, epigenetic modulators that restore normal methylation patterns at the S100B gene locus could reverse persistent overexpression even after the resolution of inflammation. For high-risk patient such as those with prolonged inflammatory episodes during neurodevelopment routine surveillance with imaging tools sensitive to glial proliferation, along with S100B/RAGE molecular profiling, could offer a proactive approach to tumor risk management. Ultimately, targeting the pathological axis of S100B overexpression and RAGE-mediated inflammation could not only improve neurological recovery but also reduce the long-term risk of glial-derived tumors such as astrocytomas and gliomas.

#### 5. Hey1

##### Neuroinflammation-Mediated Dysregulation in Glial Cells

Hey1 (Hairy/enhancer-of-split related with YRPW motif 1) is a key transcriptional repressor involved in the Notch signaling pathway, which plays a critical role in gliogenesis, particularly in the regulation of astrocyte differentiation and maintenance. During normal brain development, Hey1 regulates the timing and extent of gliogenesis, helping to balance the generation of astrocytes, oligodendrocytes, and neurons from neural progenitor cells [41]. Hey1 exerts its effects by inhibiting the transcription of genes involved in neuronal differentiation and promoting the expression of astrocytic markers, which ensures proper astrocyte maturation. However, in the context of brain inflammation, Hey1 expression can become dysregulated, resulting in significant disruptions in glial cell fate and the differentiation process. In inflammatory conditions, pro-inflammatory cytokines such as IL-6, TNF-α, and IL-1β are upregulated, and these cytokines can directly influence the expression of genes involved in Notch signaling, including Hey1. Increased levels of these cytokines can lead to altered activation of the Notch pathway, which in turn affects Hey1 gene expression. Inflammatory signaling can cause persistent activation of the Notch pathway through the upregulation of its ligands, such as Jagged1 and Delta-like 1, which leads to enhanced activation of the Notch receptor and subsequent activation of downstream signaling. This sustained activation can result in the overexpression of Hey1 in neural progenitor cells, altering the normal balance of differentiation signals [42]. Hey1’s role as a transcriptional repressor can thus shift the fate of progenitor cells toward glial lineages, particularly astrocytes, at the expense of neuronal or oligodendrocyte development. This dysregulation of glial differentiation may contribute to the increased production of reactive astrocytes in response to inflammation. Additionally, the inflammatory milieu may alter the epigenetic landscape of Hey1, leading to changes in chromatin accessibility and modifications at the Hey1 promoter region. For example, increased acetylation or phosphorylation of histones, induced by inflammation, can lead to enhanced expression of Hey1, even in the absence of strong Notch pathway activation. Furthermore, inflammatory signaling can modify the activity of other signaling pathways, such as the MAPK and JAK-STAT pathways, which can indirectly influence Hey1 expression. These pathway alterations may further exacerbate the dysregulated glial cell differentiation that occurs in response to inflammation [43]. The dysregulation of Hey1 due to brain inflammation can have long-term consequences on glial cell fate and brain function. In the short term, it leads to the abnormal accumulation of reactive astrocytes, which may alter the normal structure and function of the brain. Over time, this persistent inflammatory environment, along with the ongoing dysregulation of Hey1, may predispose to a failure in normal glial maturation and could contribute to the pathogenesis of various neurodegenerative diseases or gliomas. In summary, brain inflammation can induce a cascade of signaling events that dysregulate the expression of Hey1, which in turn disrupts the normal developmental processes of gliogenesis, contributing to pathological outcomes such as reactive gliosis and an increased susceptibility to brain tumor formation [44].

##### Consequences for Glial Cell Fate and Long-Term Brain Tumor Risk

Brain inflammation has a profound impact on the expression and function of Hey1, a critical transcriptional repressor in the Notch signaling pathway, which regulates glial cell differentiation and maturation. Under normal conditions, Hey1 controls the balance between neurogenesis and gliogenesis by promoting the differentiation of neural progenitors into astrocytes while inhibiting neuronal fate. However, during brain inflammation, the inflammatory cytokines such as IL-6, TNF-α, and IL-1β are upregulated, leading to altered Notch signaling. Inflammatory signaling may result in the sustained activation of the Notch pathway, further increasing the expression of Hey1. This dysregulation of Hey1 expression disrupts the normal differentiation of glial progenitors, leading to an excess of reactive astrocytes and impaired maturation of oligodendrocytes and neurons [45]. The elevated levels of Hey1, driven by inflammation, not only skew the differentiation towards astrocytes but also increase the production of reactive glial cells that are less supportive of neural functions. This shift in the glial cell population alters the structural and functional integrity of the brain, as reactive astrocytes, though initially protective, can become harmful when they persist and fail to resolve the inflammatory response. Over time, these aberrantly differentiated glial cells can accumulate genetic and epigenetic changes that destabilize their normal functions. These dysregulated astrocytes are more likely to acquire mutations due to the chronic inflammatory environment, where oxidative stress, cytokine signaling, and DNA damage accumulate over time. This cellular instability creates an environment conducive to the transformation of glial cells into tumorigenic phenotypes. In the long term, the continued dysregulation of Hey1 expression in the setting of chronic inflammation can foster an environment where glial progenitors and reactive astrocytes are increasingly prone to oncogenic transformations. Persistent activation of the Notch pathway through overexpression of Hey1 may contribute to the evasion of normal growth control mechanisms, allowing for unrestrained glial cell proliferation. Moreover, this dysregulated signaling can inhibit apoptosis and promote the survival of mutated cells, thus allowing them to expand into a neoplastic population [46]. The loss of proper glial differentiation and the expansion of immature, proliferative glial cells, exacerbated by chronic inflammatory signaling, creates the cellular context in which gliomas and other brain tumors may arise. Furthermore, the inflammatory-induced epigenetic modifications at the Hey1 locus, which can alter chromatin accessibility and transcriptional regulation, may lock glial progenitors into a state of sustained activation of the Notch pathway, promoting the maintenance of an undifferentiated, proliferative phenotype. This dysregulation of Hey1 not only disturbs the developmental trajectory of glial cells but also sets the stage for malignant transformation, as the continuous cycles of inflammation, abnormal cell fate decisions, and unchecked proliferation increase the risk of glioma formation.

Thus, brain inflammation’s impact on Hey1 significantly disrupts glial cell fate, leading to an increased risk of brain tumors later in life, particularly gliomas, by promoting an inflammatory environment that drives aberrant glial differentiation and facilitates tumorigenic transformations [47, 48].

##### Possible Clinical Implications in Patients: Toward Improved Diagnosis, Surveillance, and Therapeutics

The inflammation-induced dysregulation of Hey1, a critical transcriptional repressor in the Notch signaling pathway, has substantial clinical implications for improving diagnostic accuracy and long-term patient monitoring following neuroinflammatory insults. Hey1 is essential for the controlled differentiation of neural progenitors into astrocytes, and its overexpression during brain inflammation, driven by cytokines such as IL-6 and TNF-α, can skew gliogenesis toward reactive astrocytosis. Clinically, abnormal elevations of Hey1 or its upstream Notch signaling components may serve as early molecular indicators of glial imbalance in patients recovering from encephalitis, meningitis, or traumatic brain injury. Detecting such dysregulation through CSF biomarkers, gene expression profiling, or neuroimaging of reactive gliosis may help clinicians identify patients at elevated risk for long-term glial dysfunction or neoplastic transformation. Early identification of sustained Hey1 overactivity could also guide risk-based stratification and follow-up, especially in pediatric populations where glial plasticity is highest and vulnerability to inflammatory insults is most pronounced. Therapeutically, targeting the Hey1-Notch axis offers potential for intervention in both the acute and chronic phases of CNS inflammation. Notch pathway inhibitors, small interfering RNAs against Hey1, or epigenetic modulators that restore normal transcriptional regulation of Hey1 may help rebalance astrocyte differentiation and mitigate the expansion of reactive glial cells. In cases where persistent Hey1 activation is linked with impaired neuronal or oligodendrocyte development, combinatorial strategies that support neurogenesis or remyelination may be necessary. Furthermore, ongoing surveillance in high-risk patients could involve periodic molecular assessments of Hey1 expression and Notch activity to detect early signs of tumorigenic glial behavior. Since chronic overactivation of Hey1 is implicated in the formation of gliomas through mechanisms such as suppression of apoptosis, unchecked proliferation, and impaired glial maturation, modulating this pathway may not only restore developmental balance but also serve as a preventive measure against inflammation-associated brain tumors. Ultimately, understanding and targeting Hey1 dysregulation represents a critical step toward more personalized and proactive approaches in neuro-oncology and neuroinflammatory disease management.

#### 6. HES1

##### Neuroinflammation-Mediated Dysregulation in Glial Cells

HES1 (Hairy and Enhancer of Split 1) is a key transcriptional repressor in the Notch signaling pathway, playing a crucial role in regulating gliogenesis by controlling the differentiation of neural progenitors into glial cells. During normal development, HES1 acts to maintain the undifferentiated state of neural progenitors and regulates the timing of gliogenesis by repressing the expression of genes involved in astrocyte differentiation. In the presence of inflammatory signals, however, the expression and activity of HES1 may become dysregulated, leading to alterations in glial cell fate decisions and promoting pathological states in the brain. Brain inflammation, driven by the release of pro-inflammatory cytokines such as IL-1β, TNF-α, and IL-6, is known to activate various signaling pathways that intersect with Notch signaling, including the JAK-STAT and NF-κB pathways [49]. These inflammatory mediators can disrupt the normal regulation of HES1, causing abnormal activation or repression of its expression. In inflammatory conditions, the pro-inflammatory cytokines can enhance the activation of Notch receptors on neural progenitor cells, leading to a sustained activation of downstream targets such as HES1. This persistent activation of the Notch signaling pathway, in combination with the inflammatory signals, may result in prolonged HES1 expression, which can delay or inhibit the differentiation of glial cells. Under normal circumstances, HES1 serves to balance the timing of gliogenesis, but when dysregulated by inflammation, it can block the differentiation of astrocytes and other glial cell types. This sustained repression of glial differentiation can lead to a reduced number of functional glial cells, particularly astrocytes, which are essential for maintaining brain homeostasis, protecting neurons, and supporting neurogenesis. The inability of progenitor cells to properly differentiate into glial cells can thus result in an imbalance in the glial population and contribute to neuroinflammation, gliosis, and other pathological states. Furthermore, inflammation-induced changes in the epigenetic regulation of HES1 may contribute to the dysregulation of its expression [50].

Chronic inflammation often leads to altered chromatin modifications, such as histone acetylation or DNA methylation, which can impact the accessibility of the HES1 promoter region. These epigenetic modifications may either enhance or suppress HES1 expression, depending on the specific inflammatory signals present. In addition, other signaling pathways activated during brain inflammation, such as the MAPK and PI3K pathways, can cross-talk with the Notch signaling pathway, further influencing HES1 expression and activity. This complex regulatory network makes it difficult for neural progenitors to maintain their normal differentiation program and increases the potential for cellular fate abnormalities. The dysregulated expression of HES1 in the context of brain inflammation can also have long-term consequences. Prolonged activation of HES1 in neural progenitors can result in the accumulation of undifferentiated progenitor cells or reactive glial cells, which are more prone to genetic and epigenetic changes [51]. These changes can predispose the cells to oncogenic transformations, as inflammation-driven alterations in gene expression, coupled with cellular proliferation, create an environment ripe for tumorigenesis. Dysregulated HES1 activity, in combination with other inflammatory and genetic insults, can push neural progenitor cells toward abnormal proliferative and survival pathways, setting the stage for the development of gliomas or other brain tumors later in life. Thus, brain inflammation’s impact on HES1 expression disrupts the delicate balance required for proper glial differentiation, increasing the risk of neurodegenerative diseases and brain tumors through a combination of cellular misdifferentiation, abnormal proliferation, and the accumulation of genetic damage [52].

##### Consequences for Glial Cell Fate and Long-Term Brain Tumor Risk

Brain inflammation can have a significant impact on the expression and function of HES1, a transcriptional repressor involved in the Notch signaling pathway, which is crucial for regulating glial cell differentiation during development. Under normal conditions, HES1 helps control the timing of gliogenesis by inhibiting the premature differentiation of neural progenitors into glial cells. However, when the brain experiences inflammation due to factors such as injury, infection, or chronic diseases, inflammatory cytokines such as IL-6, TNF-α, and IL-1β can disrupt the normal regulation of Notch signaling and lead to aberrant activation or repression of HES1 expression [53]. Inflammatory signals often upregulate Notch receptor activation, which in turn can increase HES1 expression, keeping neural progenitors in an undifferentiated or proliferative state for longer periods. This disruption prevents the proper differentiation of glial cells, particularly astrocytes, oligodendrocytes, and microglia, which are essential for maintaining brain homeostasis and supporting neuronal health. The dysregulation of HES1 due to brain inflammation can create a prolonged inhibition of glial differentiation, leading to an accumulation of undifferentiated or poorly differentiated glial progenitors. These cells, which should normally develop into specialized glial cell types, instead remain in a proliferative or reactive state, increasing the risk of neuroinflammation and gliosis. Furthermore, this failure in proper glial differentiation can result in a functional deficit of mature astrocytes, which play key roles in neuroprotection, synaptic maintenance, and repair after injury. As a consequence, the brain’s capacity to resolve inflammation and restore homeostasis is compromised, creating an environment conducive to further damage and dysfunction. In addition to altering the differentiation of glial cells, the prolonged activation of HES1 due to inflammation can promote a cellular environment that is prone to oncogenic transformation [54]. The accumulation of undifferentiated progenitor cells, combined with the chronic inflammatory environment, may lead to the accumulation of genetic and epigenetic changes that increase the potential for malignant transformation. This is because inflammation-induced oxidative stress and DNA damage can accumulate in these reactive or proliferative glial cells, which are less capable of handling the stress of an inflamed microenvironment. The ongoing cell proliferation in the absence of proper differentiation signals can increase the likelihood of genetic mutations, particularly in genes associated with cell cycle regulation, apoptosis, and tumor suppression. Additionally, the chronic dysregulation of HES1 and its impact on glial cell fate decisions can lead to a loss of growth control mechanisms in neural progenitors. With HES1’s role in preventing excessive differentiation, its prolonged activity in an inflammatory setting can drive the survival of cells that would otherwise undergo apoptosis or senescence, thus promoting the persistence of potentially tumorigenic cells. These persistent, undifferentiated cells are at a heightened risk of accumulating mutations that enable them to bypass normal growth control checkpoints, leading to the development of gliomas or other brain tumors later in life [55]. Over time, the combined effects of chronic inflammation, sustained Notch signaling, and dysregulated HES1 expression create an environment where glial progenitors can acquire the necessary traits for tumorigenesis, including unchecked proliferation, evasion of cell death, and altered differentiation pathways. Thus, brain inflammation’s impact on HES1 not only disrupts the normal cell fate of glial cells but also lays the groundwork for the development of brain tumors by promoting an environment conducive to genetic instability, tumor formation, and the persistence of undifferentiated proliferative cells [56].

##### Possible Clinical Implications in Patients: Toward Improved Diagnosis, Surveillance, and Therapeutics

The inflammation-driven dysregulation of HES1 in neural progenitors can delay or impair glial cell differentiation, resulting in an accumulation of undifferentiated or reactive glial cells that contribute to gliosis and neuroinflammation. Clinically, this may hinder effective brain repair, exacerbate neurodegenerative processes, and increase long-term tumorigenic risk, particularly gliomas. Recognizing HES1 as a molecular mediator of inflammation-induced gliogenic disruption provides a promising avenue for early diagnostic markers and risk stratification tools.

Monitoring HES1 expression could serve as a biomarker for identifying patients at higher risk of glial dysregulation or malignant transformation. Therapeutically, targeting upstream inflammatory signals or modulating Notch-HES1 activity could offer precision strategies to restore glial cell balance, improve neuroregeneration, and prevent progression to brain tumors.

#### 7. DTX

##### Neuroinflammation-Mediated Dysregulation in Glial Cells

DTX (Deltex) proteins are part of a family of E3 ligases involved in the regulation of the Notch signaling pathway, which is essential for the proper development of glial cells during gliogenesis. In the context of gliogenesis, DTX proteins act as modulators of the Notch signaling cascade, which regulates the balance between neural progenitor cell proliferation and differentiation into various glial cell types, including astrocytes and oligodendrocytes. Under normal developmental conditions, DTX proteins help fine-tune the activation of Notch receptors, ensuring that neural progenitors differentiate at the appropriate time and in a controlled manner. However, during brain inflammation, which can be triggered by injury, infection, or chronic neurodegenerative diseases, inflammatory cytokines such as IL-1β, TNF-α, and IL-6 can disrupt the normal function of Notch signaling and, consequently, alter the expression and activity of DTX proteins [57]. Inflammatory cytokines and other signaling molecules induced during brain inflammation can affect the expression of DTX genes either by upregulating or downregulating their activity. In particular, inflammatory signals such as TNF-α and IL-6 are known to activate NF-κB and JAK-STAT pathways, both of which can influence the transcriptional activity of Notch-related genes, including DTX. These inflammatory signals can potentially enhance or diminish DTX expression, leading to an imbalance in the regulation of Notch signaling. When DTX activity is dysregulated due to inflammation, it can lead to aberrant Notch signaling, which in turn may prevent the proper differentiation of neural progenitors into glial cells [58]. Overactivation of Notch signaling due to excessive DTX activity may result in an expansion of undifferentiated progenitors and a delay in glial differentiation. On the other hand, reduced DTX expression could lead to insufficient Notch activation, impairing the maintenance of progenitor cells and reducing the pool of glial precursors available for differentiation. The impact of brain inflammation on DTX expression can also lead to other downstream consequences. For instance, dysregulated Notch signaling, influenced by altered DTX activity, can disrupt the balance of progenitor cell proliferation and differentiation, potentially resulting in the accumulation of abnormal glial cell types. This imbalance could lead to gliosis, a reactive process where glial cells proliferate in response to injury or inflammation, but without the normal regulatory mechanisms in place. Gliosis may result in the formation of reactive astrocytes and oligodendrocytes that are functionally impaired, contributing to neuroinflammation and tissue damage. In the long term, these abnormal glial cells may become more susceptible to genetic mutations and epigenetic modifications, further increasing the risk of gliomas and other brain tumors. Moreover, the inflammatory milieu may also impact the epigenetic regulation of DTX genes, as inflammatory cytokines can alter chromatin structure and DNA methylation patterns [59]. These changes in the epigenome may cause persistent alterations in DTX gene expression, even after the inflammatory response subsides. Chronic activation of Notch signaling due to persistent DTX dysregulation can increase cellular proliferation and survival, creating a favorable environment for oncogenic transformations. Over time, the sustained presence of undifferentiated progenitor cells, combined with the accumulation of genetic and epigenetic alterations, could lead to the formation of tumors. Furthermore, inflammation-induced oxidative stress and DNA damage may increase the mutational burden in progenitor cells, further promoting tumorigenesis. In summary, brain inflammation can disrupt the normal regulation of DTX proteins, leading to aberrant Notch signaling that affects gliogenesis. This dysregulation may prevent proper glial differentiation, contributing to neuroinflammation and gliosis. Moreover, chronic inflammation and altered DTX expression can promote the accumulation of undifferentiated or abnormal glial cells, which may increase the risk of brain tumors later in life due to genetic instability, sustained cell proliferation, and the accumulation of mutations [60]

##### Consequences for Glial Cell Fate and Long-Term Brain Tumor Risk

Brain inflammation can profoundly affect the expression and function of DTX (Deltex) proteins, which play a crucial role in modulating the Notch signaling pathway during gliogenesis. DTX proteins function as E3 ligases that regulate Notch receptor activation, a signaling cascade that is essential for controlling the balance between neural progenitor cell proliferation and differentiation into glial cell types such as astrocytes, oligodendrocytes, and microglia. Under normal conditions, DTX proteins help ensure that Notch signaling is appropriately activated, preventing premature differentiation of progenitor cells and allowing for the controlled development of glial cells. However, during brain inflammation, which may occur due to infection, injury, or chronic neurodegenerative conditions, inflammatory cytokines such as IL-6, TNF-α, and IL-1β can disrupt the normal regulatory mechanisms of Notch signaling [61]. These cytokines activate several signaling pathways, including NF-κB and JAK-STAT, which can alter the expression of DTX proteins, either upregulating or downregulating their activity. This dysregulation of DTX expression in response to brain inflammation can have significant effects on the fate of glial cells. If DTX activity is excessively upregulated due to inflammatory signaling, it may result in prolonged activation of Notch signaling, which could inhibit the differentiation of neural progenitors into glial cells. This failure to differentiate could lead to an accumulation of undifferentiated or reactive progenitor cells in the brain, thus disrupting normal glial development and function. On the other hand, if DTX expression is reduced or impaired due to inflammatory conditions, the Notch pathway may not be sufficiently activated, impairing the maintenance of progenitor cell populations and reducing the availability of glial precursors. This disruption in the balance of progenitor cell proliferation and differentiation may lead to a reduction in mature glial cells, thereby compromising the structural and functional integrity of the brain. In the long term, these disturbances in glial cell fate decisions could have severe consequences for brain health. The accumulation of undifferentiated or reactive glial cells, especially in the context of chronic inflammation, can result in gliosis, a process where glial cells proliferate in response to injury or damage but without the normal regulatory controls in place [62].

This pathological glial proliferation can cause neuroinflammation, disrupt tissue homeostasis, and contribute to neurodegenerative processes. Additionally, this environment of persistent inflammation and aberrant glial cell activity can increase the risk of oncogenic transformations. The prolonged activation of progenitor cells, which are less differentiated and more prone to genetic instability, creates an environment where mutations may accumulate over time, further heightening the potential for the development of brain tumors. Chronic inflammation is a well-known risk factor for tumorigenesis, as it can induce DNA damage, promote oxidative stress, and disrupt normal cell cycle regulation. In the case of glial cells, this can lead to the malignant transformation of undifferentiated progenitors, resulting in the development of gliomas or other forms of brain tumors. Moreover, the dysregulation of DTX expression during brain inflammation may exacerbate these risks by preventing the proper regulation of Notch signaling and, consequently, impairing the mechanisms that control glial differentiation and apoptosis. In the absence of appropriate differentiation signals, glial progenitors may bypass normal growth control mechanisms, leading to uncontrolled cell proliferation [63]. This, combined with the inflammatory microenvironment, could result in the emergence of tumorigenic glial cells that possess enhanced survival capabilities, reduced sensitivity to apoptotic signals, and an increased ability to undergo neoplastic transformation. Over time, the accumulation of such transformed cells could form the basis for gliomas or other types of brain tumors. In conclusion, brain inflammation can significantly impact the expression and function of DTX proteins, disrupting the normal regulation of Notch signaling and glial cell differentiation. This dysregulation may lead to the accumulation of undifferentiated or reactive progenitor cells, contributing to gliosis and neuroinflammation. Moreover, the chronic inflammatory environment, coupled with the aberrant cell fate decisions of glial progenitors, can increase the likelihood of genetic mutations and tumorigenic transformations, ultimately elevating the risk of brain tumors later in life [64].

##### Possible Clinical Implications in Patients: Toward Improved Diagnosis, Surveillance, and Therapeutics

The inflammation-induced dysregulation of DTX (Deltex) proteins, which fine-tune Notch signaling during gliogenesis, presents critical clinical implications for improving diagnostic vigilance and long-term neuro-oncological surveillance in patients with neuroinflammatory conditions. Aberrant DTX expression, either upregulated or suppressed by inflammatory cytokines like IL-6 and TNF-α, can lead to improper timing of glial differentiation, resulting in an accumulation of undifferentiated or reactive glial progenitors. Clinically, identifying elevated or unstable levels of DTX and associated Notch pathway components in cerebrospinal fluid or through transcriptomic profiling could help flag patients at risk for glial dysfunction or delayed neurodevelopment. This is particularly relevant in pediatric or post-infectious populations where Notch signaling precision is essential. Furthermore, surveillance imaging combined with molecular markers of DTX activity may provide early warnings of pathological gliosis or pre-neoplastic glial proliferation. Therapeutically, modulating DTX expression or restoring controlled Notch signaling through selective pathway inhibitors or epigenetic modulators could help prevent glial overproliferation and promote proper maturation. In patients with chronic or relapsing neuroinflammation, these strategies could mitigate long-term risks such as glioma formation by preventing the persistence of progenitor pools vulnerable to mutation and transformation in an inflammatory microenvironment.

#### 8. IL-6

##### Neuroinflammation-Mediated Dysregulation in Glial Cells

IL-6 (Interleukin-6) is a multifunctional cytokine that plays a significant role in the regulation of gliogenesis, influencing the differentiation, proliferation, and survival of glial cells during brain development. Under normal conditions, IL-6 contributes to the maintenance of the neural stem cell pool and the differentiation of these stem cells into various glial cell types, including astrocytes, oligodendrocytes, and microglia. IL-6 activates its receptor complex, consisting of the IL-6 receptor (IL-6R) and the signal-transducing subunit glycoprotein 130 (gp130), which subsequently activates intracellular signaling pathways, most notably the JAK-STAT pathway. The activation of STAT3 by IL-6 is particularly important in gliogenesis, as it regulates the transcription of genes involved in cell survival, proliferation, and differentiation [65]. Additionally, IL-6 is known to play a role in reactive gliosis, a process where glial cells, particularly astrocytes, undergo hypertrophy and proliferate in response to brain injury or inflammation. In the context of brain inflammation, the expression of IL-6 is significantly upregulated due to the activation of immune responses in response to injury, infection, or chronic neurodegenerative conditions. Inflammatory mediators such as TNF-α, IL-1β, and interferons can induce the production of IL-6, leading to a cytokine cascade that amplifies inflammation in the brain. This upregulation of IL-6 during inflammation can disrupt normal gliogenesis in several ways. Firstly, prolonged or excessive IL-6 signaling can lead to the sustained activation of STAT3, which, while important for glial differentiation in moderation, may promote the differentiation of glial cells at inappropriate times or in an uncontrolled manner during chronic inflammation [66]. This can lead to an imbalance in the generation of specific glial cell types, impairing the formation of mature and functional glial networks. Overactivation of STAT3, for example, has been shown to promote astrogliosis, a pathological state where astrocytes proliferate excessively in response to injury, potentially leading to the formation of reactive glial scars that interfere with tissue repair and regeneration.

Moreover, dysregulation of IL-6 signaling in the context of chronic brain inflammation can result in an abnormal activation of downstream signaling pathways, including the MAPK and PI3K/Akt pathways, which are involved in cell survival, proliferation, and migration. These pathways can further contribute to the development of reactive gliosis and interfere with normal glial differentiation [67]. Chronic inflammation can also lead to a shift in the balance of pro-inflammatory and anti-inflammatory cytokines, exacerbating the effects of IL-6 signaling. In the case of IL-6 overproduction, this dysregulation can skew glial progenitor cells toward aberrant differentiation or even lead to the accumulation of undifferentiated progenitors that are more prone to neoplastic transformation. In the long term, the dysregulation of IL-6 signaling due to brain inflammation can create a cellular environment conducive to tumorigenesis. The persistence of IL-6 signaling in the inflamed brain can lead to the sustained activation of pathways that promote cell survival and proliferation, such as the PI3K/Akt and MAPK pathways. These pathways, when continuously activated, can increase the likelihood of mutations and genomic instability in glial cells, which may contribute to the development of gliomas or other forms of brain tumors. Additionally, the chronic activation of IL-6 and its downstream pathways, particularly STAT3, has been implicated in the inhibition of apoptosis and the promotion of cell cycle progression, both of which are key factors in tumorigenesis. The accumulation of genetically altered glial progenitors or reactive astrocytes in response to persistent IL-6 signaling may provide a substrate for tumorigenic transformations, further increasing the risk of brain tumors later in life. In conclusion, brain inflammation can disrupt the normal regulatory roles of IL-6 in gliogenesis, leading to the dysregulation of glial cell differentiation, proliferation, and survival. The upregulation of IL-6 during inflammation can result in the prolonged activation of STAT3 and other downstream signaling pathways, contributing to the pathological overproduction of glial cells, particularly astrocytes, and impairing the balance of glial cell differentiation [68].

##### Consequences for Glial Cell Fate and Long-Term Brain Tumor Risk

Brain inflammation significantly impacts IL-6 (Interleukin-6) expression, which plays a crucial role in regulating glial cell fate and function. IL-6 is a key cytokine that mediates the inflammatory response in the central nervous system (CNS) and is involved in glial differentiation, survival, and proliferation. Under normal conditions, IL-6 is involved in maintaining a balance between glial progenitor cell proliferation and differentiation, promoting proper gliogenesis. However, during brain inflammation, such as in response to injury, infection, or neurodegenerative diseases, the production of IL-6 is markedly upregulated, leading to a cascade of signaling events that can disrupt the delicate balance of glial cell fate [69]. This overproduction of IL-6 activates several intracellular signaling pathways, notably the JAK-STAT pathway, which regulates key processes involved in cell survival, proliferation, and differentiation. In particular, the activation of STAT3, a key transcription factor downstream of IL-6, can have both protective and detrimental effects on glial cell fate, depending on the duration and context of its activation. In the setting of chronic brain inflammation, excessive or prolonged IL-6 signaling can lead to aberrant STAT3 activation. STAT3 is essential for the differentiation of glial cells, including astrocytes, oligodendrocytes, and microglia, but its uncontrolled activation can drive excessive glial cell proliferation and inhibit the proper differentiation of progenitor cells. This can result in an imbalance in glial cell populations, with an overabundance of reactive astrocytes or undifferentiated glial progenitors that fail to mature into functional cells. Reactive gliosis, a pathological state characterized by the overproduction of astrocytes in response to injury or chronic inflammation, is a direct consequence of dysregulated IL-6 signaling. In this state, astrocytes undergo hypertrophy and hyperplasia, potentially forming a glial scar that impedes tissue regeneration and disrupts neuronal function [70]. The continued presence of an inflammatory microenvironment driven by IL-6 signaling may exacerbate this pathological glial response, further hindering proper brain function and repair mechanisms. Moreover, the chronic activation of IL-6 signaling in the brain’s inflammatory environment increases the risk of genetic instability and malignant transformation of glial cells. IL-6 signaling can activate survival pathways, such as the PI3K/Akt and MAPK pathways, that promote cell proliferation and inhibit apoptosis. These pathways, when persistently activated in glial progenitors or reactive glial cells, may lead to the accumulation of mutations and genomic instability. Such cellular changes can predispose glial cells to neoplastic transformation. Glial tumors, particularly gliomas, are commonly associated with dysregulated growth factors and inflammatory signals that drive the uncontrolled proliferation of glial cells. Inflammatory cytokines like IL-6 can promote an oncogenic environment where the normal regulatory mechanisms governing cell cycle progression and apoptosis are disrupted, thereby facilitating the development of tumors. Furthermore, the sustained activation of STAT3 in response to IL-6 has been implicated in the inhibition of apoptotic pathways and the promotion of cell survival, both of which are key drivers of tumorigenesis. As the inflammatory process continues, the persistent upregulation of IL-6 can contribute to the selection of glial cells that have acquired oncogenic properties. The accumulation of mutated or genetically unstable glial progenitors or reactive astrocytes increases the likelihood of these cells undergoing transformation into malignant tumors. The chronic inflammatory state, in conjunction with the dysregulated IL-6 signaling, creates a permissive environment for tumor initiation and progression. Gliomas and other brain tumors often arise from these aberrant glial cells, which proliferate uncontrollably and fail to undergo normal differentiation [71]. Moreover, the inflammatory milieu driven by IL-6 can support the survival of these transformed cells by promoting angiogenesis, immune evasion, and resistance to chemotherapeutic agents. In summary, brain inflammation has a significant impact on IL-6 signaling, disrupting the normal fate of glial cells. The excessive activation of IL-6 and its downstream pathways, particularly STAT3, can lead to uncontrolled glial proliferation and the formation of reactive glial scars, hindering proper brain function and repair. Chronic inflammation further contributes to genomic instability and the increased risk of malignant transformations in glial cells, increasing the likelihood of brain tumors, such as gliomas, later in life. The persistence of inflammatory cytokines like IL-6, coupled with the disruption of normal glial differentiation, creates a tumorigenic environment that facilitates the development and progression of brain tumors [72].

##### Possible Clinical Implications in Patients: Toward Improved Diagnosis, Surveillance, and Therapeutics

The dysregulation of IL-6 signaling during neuroinflammation presents critical opportunities for improving clinical diagnosis, surveillance, and targeted therapeutics in patients vulnerable to glial dysfunction and tumorigenesis. Elevated IL-6 levels, particularly in cerebrospinal fluid or blood, may serve as a valuable biomarker for early detection of pathological glial activation and reactive gliosis following CNS infections, trauma, or autoimmune conditions.

Clinically, persistent IL-6 elevation could be used to identify patients at higher risk for developing glial scars, neurodevelopmental disruption, or tumor-prone cellular states. Monitoring IL-6 activity alongside STAT3 phosphorylation or PI3K/Akt activation may enhance risk stratification in pediatric or chronically inflamed patient populations. This surveillance can help detect aberrant glial dynamics before the onset of irreversible neurodegeneration or malignancy. Therapeutically, modulating IL-6 signaling may help prevent the transition from adaptive gliogenesis to pathological gliosis and reduce the long-term risk of glioma formation. Pharmacologic agents such as IL-6 receptor antagonists (e.g., tocilizumab) or STAT3 inhibitors could be explored to prevent the persistent activation of survival and proliferation pathways in glial progenitors. Additionally, targeting IL-6-driven pathways like PI3K/Akt or MAPK could help rebalance glial proliferation and differentiation, preserving normal neuro-glial architecture. Integrating IL-6 signaling profiles into personalized care pathways may also guide post-inflammatory management strategies, allowing clinicians to intervene before glial cells acquire oncogenic mutations or become resistant to apoptotic signaling. Ultimately, the recognition of IL-6 as both a driver of glial dysregulation and a contributor to tumor-permissive environments reinforces its role as a central therapeutic target in the prevention of inflammation-associated brain tumors and the preservation of long-term neurological health.

#### 9. NF-κB

##### Neuroinflammation-Mediated Dysregulation in Glial Cells

NF-κB (Nuclear Factor kappa-light-chain-enhancer of activated B cells) plays a critical role in regulating inflammation, cell survival, and immune responses. During gliogenesis, NF-κB signaling contributes to the differentiation, proliferation, and survival of glial cells. Under normal physiological conditions, NF-κB activity is tightly regulated and transient, allowing proper glial development while maintaining a balance between activation and inhibition. However, brain inflammation can lead to dysregulation of NF-κB signaling, resulting in either excessive activation or impaired inhibition of this pathway, which disrupts glial cell fate decisions and contributes to pathological outcomes [73]. The activation of NF-κB is typically triggered by inflammatory cytokines, such as TNF-α and IL-1β, or through microglial activation in response to injury or infection. This inflammatory signaling can lead to the sustained or constitutive activation of NF-κB, which in turn drives the expression of pro-inflammatory genes. This chronic activation of NF-κB signaling during inflammation can cause several downstream effects that affect gliogenesis. NF-κB signaling regulates the expression of key genes involved in cell survival and differentiation, such as those encoding growth factors and pro-survival proteins. In response to prolonged inflammation, the persistent activation of NF-κB in glial progenitor cells can inhibit their differentiation into mature glial subtypes, such as oligodendrocytes and astrocytes, and promote the maintenance of a progenitor or reactive state. This dysregulation of differentiation impairs normal gliogenesis, leading to an overabundance of undifferentiated glial progenitors and reactive glial cells that fail to perform their physiological functions effectively [74]. Additionally, prolonged NF-κB activation in glial cells during brain inflammation can exacerbate the production of reactive oxygen species (ROS) and inflammatory mediators, further perpetuating the inflammatory cycle. This heightened inflammatory environment can hinder the resolution of inflammation, leading to sustained activation of glial cells, such as astrocytes and microglia. These activated glial cells contribute to the formation of a glial scar, a pathological hallmark of chronic inflammation, which impedes tissue regeneration and disrupts neuronal function. The persistence of activated microglia and reactive astrocytes, driven by aberrant NF-κB signaling, results in a hostile environment for neuronal repair and neurogenesis, which can further impair normal brain function and repair mechanisms. Beyond its role in promoting chronic inflammation, dysregulated NF-κB signaling can also contribute to an increased risk of tumorigenesis. Aberrant activation of NF-κB has been implicated in the development and progression of various types of cancer, including gliomas. In the context of gliogenesis, excessive NF-κB activation in glial progenitor cells or reactive glial cells can promote tumorigenesis by enhancing cell survival and proliferation while inhibiting apoptosis. Inflammatory cytokines such as TNF-α and IL-1β, which are upregulated during brain inflammation, can activate NF-κB signaling and support the survival of genetically unstable glial cells [75]. This chronic activation of NF-κB may lead to the accumulation of genetic mutations, as it can suppress the activation of tumor suppressor pathways and DNA repair mechanisms. Furthermore, NF-κB-mediated transcriptional regulation of pro-survival genes, such as BCL-2 and IAPs (Inhibitor of Apoptosis Proteins), contributes to the resistance of glial cells to apoptosis, allowing abnormal cells to survive and proliferate in an inflammatory environment. The role of NF-κB in promoting angiogenesis also has important implications for tumor development. In response to inflammation, NF-κB signaling can increase the expression of angiogenic factors such as VEGF (Vascular Endothelial Growth Factor), which supports the formation of new blood vessels. This is particularly relevant in the context of glioma, where the growth of blood vessels is essential for tumor expansion. By enhancing angiogenesis, NF-κB facilitates the supply of nutrients and oxygen to growing tumors, further promoting their growth and metastasis. Additionally, the inflammatory environment created by dysregulated NF-κB signaling can support immune evasion by the tumor, further enhancing its malignancy. In summary, brain inflammation disrupts the normal regulation of NF-κB signaling, leading to sustained activation of this pathway in glial cells. This dysregulation can impair the differentiation of glial progenitors, promote reactive gliosis, and hinder tissue repair. Furthermore, the persistent activation of NF-κB in response to chronic inflammation can drive cell survival, proliferation, and resistance to apoptosis in glial cells, creating a permissive environment for the development of brain tumors [76].

##### Consequences for Glial Cell Fate and Long-Term Brain Tumor Risk

Brain inflammation can significantly impact the activity of NF-κB (Nuclear Factor kappa-light-chain-enhancer of activated B cells), a critical transcription factor involved in regulating immune responses, inflammation, and cell survival. Under normal conditions, NF-κB activation is transient and tightly controlled, facilitating proper glial cell function and differentiation. However, during brain inflammation, this pathway can become dysregulated, leading to sustained NF-κB activation [77]. Chronic activation of NF-κB in glial cells, particularly microglia and astrocytes, can disrupt the normal balance between cell proliferation, differentiation, and survival, thus impairing gliogenesis and promoting an abnormal glial cell phenotype. In the context of brain inflammation, NF-κB is often activated by cytokines such as TNF-α, IL-1β, and other inflammatory mediators that are upregulated during injury, infection, or other forms of neuroinflammation. This activation results in the expression of genes that promote cell survival and inflammation [78]. In glial cells, this pro-inflammatory signaling pathway can lead to a reactive gliosis phenotype, where glial progenitor cells and mature glial cells become activated, proliferate excessively, and fail to properly differentiate into their mature forms. Instead of completing their differentiation into functional astrocytes or oligodendrocytes, these cells may remain in a proliferative or undifferentiated state. This improper differentiation disrupts the normal development of glial cells and their ability to support neuronal function, repair, and homeostasis, leaving the brain vulnerable to further damage and dysfunction. Moreover, the chronic inflammation and sustained NF-κB activation in the glial cell population can alter the cellular microenvironment by promoting oxidative stress and the production of reactive oxygen species (ROS). These inflammatory by-products can lead to cellular DNA damage, further disturbing the normal cell cycle and increasing the potential for genetic mutations. Such mutations can result in the dysregulation of critical cell cycle control genes, impairing apoptosis and allowing genetically altered glial cells to survive and proliferate uncontrollably. As a consequence, prolonged inflammatory signaling, along with the inability of glial cells to differentiate properly, sets the stage for abnormal glial cell growth, which can increase the likelihood of tumor formation. Increased NF-κB activity also plays a role in promoting angiogenesis by upregulating vascular endothelial growth factor (VEGF) and other pro-angiogenic factors. This is particularly relevant in the context of glioma development, where the formation of new blood vessels is essential for tumor growth and survival. The persistent inflammation associated with dysregulated NF-κB signaling thus provides a favorable environment for the expansion of abnormal glial cells and the vascularization of tumors. In addition, the enhanced production of inflammatory cytokines can foster a microenvironment that allows tumors to evade immune surveillance, further facilitating tumor progression. As glial cells with aberrant NF-κB signaling continue to proliferate and accumulate mutations over time, the risk of malignant transformation increases [79]. The imbalance between cell survival and differentiation, coupled with genomic instability induced by chronic inflammation, creates conditions that are conducive to the development of gliomas and other brain tumors. Moreover, the interaction between NF-κB-driven inflammation and other molecular pathways, such as the JAK-STAT pathway and p53 signaling, can further amplify the tumorigenic potential of these glial cells. The combination of sustained inflammation, cellular dysregulation, and genetic instability driven by NF-κB can thus increase the likelihood of glioma formation, particularly in individuals who experience long-term brain inflammation, whether due to chronic neuroinflammatory diseases or acute traumatic injury. In summary, brain inflammation’s impact on NF-κB signaling plays a crucial role in disrupting the differentiation and function of glial cells, contributing to the persistence of undifferentiated or reactive glial cells that may foster an environment conducive to brain tumorigenesis. The sustained activation of NF-κB during chronic inflammation promotes cell survival, proliferation, and resistance to apoptosis, while also increasing oxidative stress and genetic instability [80].

##### Possible Clinical Implications in Patients: Toward Improved Diagnosis, Surveillance, and Therapeutics

The dysregulation of NF-κB signaling during brain inflammation has important clinical implications for both the early diagnosis and long-term surveillance of glial pathology in patients exposed to CNS inflammation. Under pathological conditions, sustained NF-κB activation, triggered by cytokines like TNF-α and IL-1β leads to chronic reactive gliosis, glial progenitor arrest, and a failure of proper differentiation. Clinically, elevated NF-κB activity in cerebrospinal fluid or glial tissues could serve as a biomarker of unresolved inflammation, glial scarring, or pre-neoplastic glial expansion. Early detection of these changes could inform post-injury or post-infection monitoring strategies in high-risk groups, such as children with meningitis or patients with neurodegenerative diseases. Imaging techniques that capture gliosis, coupled with molecular profiling of NF-κB-regulated pro-inflammatory or pro-survival genes (e.g., BCL-2, VEGF), may enhance the sensitivity of clinical surveillance and help identify patients who would benefit from anti-inflammatory or anti-proliferative therapies. Therapeutically, targeting aberrant NF-κB signaling could offer substantial benefits in halting glial overproliferation and preventing long-term neurooncological outcomes. Pharmacologic inhibitors of NF-κB or its upstream activators, alongside antioxidants that reduce reactive oxygen species (ROS), may help restore balance to glial cell differentiation and reduce inflammation-driven genetic instability. These therapies could also limit the formation of glial scars, support neural repair, and prevent the accumulation of undifferentiated glial cells that are vulnerable to transformation. In patients with recurrent neuroinflammation or traumatic brain injury, regular assessment of NF-κB activity and related angiogenic or apoptotic markers could guide the use of preventative interventions aimed at reducing glioma risk. Furthermore, the interplay between NF-κB and other signaling pathways like JAK-STAT and p53 suggests that combined pathway-targeted therapies may be necessary for effective modulation. Ultimately, integrating NF-κB monitoring into routine follow-up protocols for neuroinflammatory conditions could improve neurological outcomes, reduce the risk of tumor development, and guide more personalized and preventive neuro-oncology care.

#### 10. Neuregulin-1

##### Neuroinflammation-Mediated Dysregulation in Glial Cells

Neuregulin-1 (NRG1) plays a crucial role in gliogenesis, particularly in the development and maintenance of oligodendrocytes and other glial cell types within the central nervous system. NRG1 exerts its effects through activation of the ErbB receptor family, which influences various cellular processes, including differentiation, migration, and survival of glial cells. During normal development, NRG1 signaling is tightly regulated to ensure the appropriate maturation of oligodendrocyte progenitor cells (OPCs) into mature oligodendrocytes, the cells responsible for myelination. However, during brain inflammation, the expression of NRG1 and its signaling pathways can become dysregulated, leading to alterations in glial cell fate and function [81]. In the context of brain inflammation, such as in conditions like encephalitis or neurodegenerative diseases, inflammatory cytokines like IL-1β, TNF-α, and IL-6 are often upregulated. These inflammatory signals can interfere with the normal expression of NRG1, either through direct suppression or through the activation of signaling pathways such as NF-κB and STAT3, which may downregulate the transcription of the NRG1 gene. When NRG1 expression is disrupted, it can lead to improper differentiation of glial progenitor cells, such as oligodendrocytes, as well as an impaired ability of glial cells to respond to environmental cues that are necessary for their survival and maturation [82]. Specifically, this dysregulation may hinder the differentiation of OPCs, leaving them in a proliferative or undifferentiated state, which can contribute to insufficient myelination and impaired repair of injured neural tissue. Furthermore, the chronic activation of inflammatory pathways can alter the signaling cascade downstream of NRG1, such as the ErbB receptors, which are responsible for transmitting the signals from NRG1 to the glial cells. Under inflammatory conditions, alterations in receptor expression or function can occur, potentially leading to a shift in cellular behavior. This dysregulated signaling can promote aberrant glial cell proliferation, reactive gliosis, and an overall failure to properly regulate glial differentiation and function. The disturbed NRG1-ErbB signaling also impairs the formation of the correct glial cell populations needed for tissue homeostasis and repair. For instance, an imbalance in the differentiation of oligodendrocytes and astrocytes may result, leading to the inability of the glial cells to support neuronal function adequately. In addition to altering cell differentiation, brain inflammation can also impact the ability of NRG1 signaling to regulate the survival of glial cells [83]. Under inflammatory conditions, excessive or prolonged activation of the NRG1-ErbB pathway may lead to increased survival of reactive glial cells, which can adopt a hyperproliferative and pro-inflammatory state. This abnormal glial cell proliferation and failure to differentiate properly may contribute to the pathogenesis of brain tumors. The disrupted NRG1 signaling creates an environment conducive to the accumulation of mutations in glial cells, further promoting their transformation into tumorigenic cells. Moreover, the increased survival of reactive glial cells during inflammation creates a pool of cells that, over time, may acquire additional genetic alterations, increasing the risk of glioma formation and other brain tumors. The disruption of NRG1 signaling can also affect the balance between glial and neuronal interactions [84]. NRG1 is involved in the regulation of neuron-glia communication, particularly in terms of the myelination of axons. During brain inflammation, the failure of NRG1 to properly activate its downstream signaling pathways can compromise this communication, leading to a disruption of normal neuronal function and a reduction in the ability of the central nervous system to repair itself. The resulting cellular and molecular imbalances in glial cell populations and their interactions with neurons may promote chronic neuroinflammation, which in turn can contribute to the development of brain tumors, particularly gliomas. In conclusion, brain inflammation can significantly dysregulate the expression and signaling of NRG1, leading to impaired glial cell differentiation, aberrant proliferation, and altered cell survival.

The failure of NRG1 signaling to properly guide the differentiation and maturation of glial cells, especially oligodendrocytes, can contribute to insufficient tissue repair and neurodegeneration. Additionally, the dysregulation of NRG1 can promote a microenvironment conducive to glial cell transformation, increasing the risk of brain tumor formation later in life. Thus, understanding how brain inflammation affects NRG1 signaling and glial cell fate is critical for developing strategies to prevent and treat neuroinflammatory-related diseases and brain tumors [85].

##### Consequences for Glial Cell Fate and Long-Term Brain Tumor Risk

Brain inflammation can have a profound impact on the expression and signaling of Neuregulin-1 (NRG1), which plays a pivotal role in glial cell development and the maintenance of the central nervous system. NRG1 interacts with the ErbB receptor family to influence various cellular processes essential for glial cell differentiation, proliferation, and survival. In normal development, NRG1 signaling is crucial for guiding oligodendrocyte precursor cells (OPCs) toward their differentiation into mature oligodendrocytes, which are responsible for myelination. During brain inflammation, however, the regulatory pathways governing NRG1 expression and its signaling cascade can be disrupted, leading to significant alterations in glial cell fate and the overall health of the central nervous system. Inflammatory cytokines such as IL-1β, TNF-α, and IL-6, which are elevated during neuroinflammation, can suppress or alter the expression of NRG1. These cytokines activate inflammatory signaling pathways like NF-κB and STAT3, which can reduce NRG1 transcription, thereby interfering with the proper differentiation of glial progenitors, such as OPCs, into mature glial cells [86]. This dysregulation results in an abnormal accumulation of undifferentiated glial progenitor cells, impairing normal glial maturation and leading to deficits in myelination and cellular function. Furthermore, prolonged brain inflammation can alter the downstream signaling of the ErbB receptors, which are critical for transmitting the signals from NRG1 to glial cells. Dysregulation of ErbB receptor expression or function due to inflammation may disrupt the normal cellular responses that NRG1 typically initiates. For example, improper activation of the ErbB receptors during inflammation can lead to abnormal glial cell proliferation, impairing the balance between cell growth and differentiation. This can create an environment in which glial cells fail to mature properly, contributing to the accumulation of immature glial cells and reactive gliosis. In addition to disrupting glial cell differentiation, inflammatory-induced alterations in NRG1 signaling may also impact glial cell survival, particularly under conditions of prolonged neuroinflammation. Inflammation-induced hyperactivation of the NRG1-ErbB signaling pathway may lead to the excessive survival of reactive glial cells, further promoting their proliferation [87]. This uncontrolled growth, coupled with the failure to differentiate, can create an environment conducive to tumorigenesis. Over time, the accumulation of genetic alterations in these proliferating glial cells, exacerbated by the inflammatory milieu, could increase the likelihood of malignant transformation, contributing to the development of gliomas and other types of brain tumors. The interaction between neuroinflammation and NRG1 signaling may also disrupt the communication between glial cells and neurons. NRG1 plays a role in neuron-glia signaling, particularly in the regulation of myelination and neuronal function. Inflammatory cytokines that interfere with NRG1 expression can hinder proper myelination, leading to impaired neuronal signaling and axonal function. This disruption in glial-neuronal interactions further contributes to the pathology of brain inflammation and promotes a cycle of chronic neuroinflammation. Over time, the persistent inflammatory environment, coupled with the dysfunctional NRG1 signaling, creates an optimal setting for the accumulation of mutations in glial cells. These mutations, in conjunction with the altered proliferative and survival pathways triggered by NRG1 dysregulation, increase the risk of glioma formation and other brain tumors in the long term. In conclusion, brain inflammation has a profound impact on the regulation of NRG1 expression and signaling, leading to disruptions in glial cell differentiation, proliferation, and survival. These alterations in glial cell fate, combined with the inflammatory microenvironment, increase the likelihood of glioma formation and other brain tumors later in life. The dysregulation of NRG1 signaling during inflammation creates a cycle of abnormal glial cell growth and impaired differentiation, which can promote the development of tumors [88].

##### Possible Clinical Implications in Patients: Toward Improved Diagnosis, Surveillance, and Therapeutics

The dysregulation of Neuregulin-1 (NRG1) during brain inflammation carries significant clinical implications for early diagnosis, disease monitoring, and targeted therapeutic intervention in neuroinflammatory and neuro-oncological disorders. NRG1 is essential for guiding oligodendrocyte precursor cells (OPCs) through proper differentiation and for maintaining neuron-glia interactions, particularly in processes like myelination. Inflammatory insults that suppress NRG1 expression or impair ErbB receptor signaling can stall OPC maturation, leading to demyelination and impaired CNS repair. Clinically, altered NRG1 or ErbB signaling could serve as molecular biomarkers of disrupted glial development in patients with encephalitis, multiple sclerosis, or other neuroinflammatory syndromes. Early detection of downregulated NRG1 signaling through CSF or blood-based assays could help identify patients at risk for long-term white matter damage or glial scarring. Moreover, neuroimaging techniques that assess myelination patterns could be complemented by NRG1-based molecular profiling to enhance the precision of glial pathology assessment in both acute and chronic neuroinflammatory settings. From a therapeutic perspective, restoring NRG1-ErbB signaling may offer a valuable approach to correcting glial dysfunction and preventing tumorigenic transformation in inflamed neural environments. Pharmacological agents or biologics that enhance NRG1 expression, stabilize ErbB receptor function, or selectively promote OPC differentiation could support remyelination and CNS recovery following inflammation. Conversely, in cases where excessive NRG1 activity contributes to the survival and hyperproliferation of reactive glial cells, modulating the pathway may help prevent the buildup of undifferentiated glial progenitors at risk for oncogenic transformation. Patients with persistent neuroinflammation may benefit from long-term molecular surveillance of the NRG1 signaling axis, especially if they exhibit signs of glial hyperplasia or demyelination. Ultimately, a deeper understanding of how NRG1 dysregulation contributes to the inflammation-tumor transition can inform targeted preventive strategies against gliomas and other glial-derived tumors, especially in high-risk populations such as children recovering from CNS infections or individuals with chronic inflammatory neurological diseases.

#### 11. MAPK/MEK signaling pathway

##### Neuroinflammation-Mediated Dysregulation in Glial Cells

The MAPK/MEK signaling pathway is a crucial regulator of cellular processes, including differentiation, proliferation, survival, and migration, during gliogenesis. MEK (mitogen-activated protein kinase kinase) activates the extracellular signal-regulated kinase (ERK) via phosphorylation, which then regulates the expression of genes involved in cell fate determination, particularly in glial progenitors. This pathway is essential for the proper development of oligodendrocytes, astrocytes, and other glial cells from their precursor populations [89]. During normal development, the MAPK/MEK/ERK pathway plays a pivotal role in guiding glial differentiation and maturation, as well as ensuring the proper function of differentiated glial cells in the central nervous system. However, during brain inflammation, this pathway can become dysregulated through the action of pro-inflammatory cytokines and immune cell signaling. Inflammatory conditions often lead to an increase in cytokines such as TNF-α, IL-1β, and IL-6, which can modulate the activity of the MAPK/MEK/ERK pathway. These cytokines are known to activate NF-κB and other transcription factors, which, in turn, can lead to aberrant activation or suppression of MEK and its downstream targets. In some cases, this can result in enhanced activation of the MAPK pathway, which may promote the excessive proliferation of glial progenitors and hinder their differentiation into mature glial cells. Inflammatory signaling may also induce the phosphorylation of ERK inappropriately, leading to the upregulation of genes associated with cellular survival and proliferation, while downregulating genes responsible for differentiation [90]. The dysregulated activation of MEK/ERK can also impair the normal interaction between glial progenitors and their microenvironment, disrupting the signaling required for appropriate cell fate decisions. Furthermore, brain inflammation can also affect the expression of MAPK/MEK pathway components, such as MEK1, MEK2, and various downstream targets of ERK, including transcription factors that regulate glial differentiation. Chronic inflammation can lead to prolonged activation of the MAPK/MEK/ERK pathway, which may push glial progenitors toward a more proliferative and less differentiated state. This sustained proliferative signaling can lead to an accumulation of undifferentiated glial progenitor cells, resulting in a failure to form mature oligodendrocytes and astrocytes. Additionally, the disruption of normal differentiation processes may lead to gliosis, a pathological condition characterized by the overproduction of glial cells in response to injury or inflammation. This phenomenon can be further exacerbated by the dysregulation of other signaling pathways, such as Notch or Wnt, which interact with the MAPK/MEK/ERK pathway to influence glial differentiation. In a more long-term perspective, the chronic activation of the MAPK/MEK/ERK pathway during ongoing inflammation may contribute to the development of gliomas or other types of brain tumors [91]. The continued activation of pro-proliferative signaling in glial progenitor cells, coupled with the inhibition of differentiation, creates an environment conducive to tumorigenesis. Inflammation-induced alterations in the MAPK/MEK/ERK pathway may promote the survival of abnormal, undifferentiated glial cells, providing a pool of cells that are prone to genetic mutations and malignant transformation. As these progenitors accumulate and undergo genetic alterations, the likelihood of tumor formation increases, particularly gliomas, which arise from glial progenitor cells. Furthermore, the aberrant signaling through MAPK/MEK/ERK in the context of inflammation can also enhance the invasiveness and resistance of glial cells to apoptosis, traits that are characteristic of malignant tumors. In summary, brain inflammation has the potential to disrupt the normal function of the MAPK/MEK/ERK signaling pathway in glial cells by altering the expression and activity of key pathway components. This dysregulation can lead to an imbalance in the differentiation and proliferation of glial progenitors, contributing to gliosis, abnormal glial cell accumulation, and a failure to generate mature glial cells. In the long term, the sustained activation of the MAPK/MEK/ERK pathway under inflammatory conditions can increase the risk of brain tumor formation, particularly gliomas. Understanding the mechanisms by which brain inflammation affects MAPK/MEK signaling in gliogenesis could provide valuable insights into the development of therapeutic strategies aimed at mitigating the effects of chronic neuroinflammation and preventing the development of gliomas [92].

##### Consequences for Glial Cell Fate and Long-Term Brain Tumor Risk

Brain inflammation has the potential to significantly impact the MAPK-MEK signaling pathway, which is crucial in regulating the development, differentiation, and survival of glial cells during gliogenesis. Under normal conditions, the MAPK-MEK pathway plays a central role in the differentiation of oligodendrocytes, astrocytes, and other glial progenitor cells, facilitating the transition from proliferative states to differentiated, mature glial cells. However, brain inflammation can disrupt this pathway, leading to significant changes in the cell fate of glial progenitors, which may have long-term consequences, including an increased risk of brain tumors [93]. Inflammatory signals, such as cytokines released in response to brain injury or infections, can directly influence the activity of the MAPK-MEK signaling cascade. Pro-inflammatory cytokines like TNF-α, IL-1β, and IL-6 can activate upstream receptors that promote the activation of the MEK/ERK axis, sometimes inappropriately. This abnormal activation may drive glial progenitors into a proliferative state, preventing proper differentiation. Under normal conditions, the MAPK-MEK pathway helps orchestrate the timing and balance of glial cell differentiation. However, in the context of inflammation, this signaling pathway may become hyper-activated or dysregulated, causing glial progenitors to proliferate uncontrollably and impairing their differentiation into fully functional glial cells. Instead of becoming mature astrocytes or oligodendrocytes, these progenitors may remain in an undifferentiated, proliferative state. In the presence of chronic inflammation, the sustained activation of the MAPK-MEK pathway can lead to a buildup of undifferentiated glial progenitors. These progenitors are more likely to accumulate genetic mutations over time due to the increased cell turnover and defective differentiation. The genetic instability introduced by chronic inflammation and sustained MEK/ERK signaling may contribute to the emergence of abnormal glial cell populations [94]. This may provide the cellular basis for gliomas and other types of brain tumors, which often arise from glial progenitor cells that undergo malignant transformation. The abnormal signaling also interferes with normal cell cycle regulation, reducing apoptosis and increasing the survival of these potentially transformed cells. As these altered progenitors continue to divide, the risk of tumorigenesis increases. Moreover, the dysregulation of the MAPK-MEK signaling pathway in the context of brain inflammation can alter the microenvironment surrounding glial cells.

Chronic inflammation leads to the release of additional pro-inflammatory factors, such as reactive oxygen species (ROS), which can damage DNA and further increase the likelihood of oncogenic mutations. This creates a feedback loop where inflammatory signals and MAPK-MEK pathway activation continue to support the proliferation of aberrant glial progenitors. Over time, this chronic dysregulation can lead to the expansion of abnormal glial populations, increasing the risk of malignant transformation. Furthermore, the altered activation of the MAPK-MEK pathway during brain inflammation may also affect glial cell function. Astrocytes, for example, are essential for maintaining the blood-brain barrier, providing metabolic support to neurons, and managing inflammatory responses. Disruption of proper astrocyte differentiation due to impaired MAPK-MEK signaling may lead to dysfunctional glial cells that cannot adequately support neuronal health, exacerbating the effects of ongoing inflammation. In some cases, the aberrant activation of MEK/ERK may also interfere with the maturation of oligodendrocytes, impairing myelination and promoting the development of demyelinating conditions, which may create a more permissive environment for the development of brain tumors. In summary, brain inflammation can disrupt the MAPK-MEK signaling pathway, leading to a failure of glial progenitor cells to properly differentiate into mature glial cells [95]. Chronic activation of this pathway under inflammatory conditions can result in an accumulation of undifferentiated glial progenitors that are prone to genetic instability and malignant transformation. Over time, this dysregulated signaling may increase the risk of glioma and other brain tumors by creating an environment where abnormal glial cell proliferation is favored over differentiation, and the normal regulatory mechanisms of cell death and survival are disrupted. Understanding these pathways provides valuable insights into how chronic inflammation can contribute to brain tumorigenesis, highlighting potential therapeutic targets to mitigate these risks [96].

##### Possible Clinical Implications in Patients: Toward Improved Diagnosis, Surveillance, and Therapeutics

Dysregulation of the MAPK/MEK signaling pathway during neuroinflammation presents critical clinical implications for improving diagnostic precision, long-term surveillance, and therapeutic strategies in patients at risk of glial dysfunction and brain tumors. This pathway, which normally guides glial progenitor differentiation into mature astrocytes and oligodendrocytes, becomes abnormally activated under inflammatory conditions due to cytokines like IL-1β, IL-6, and TNF-α. Such dysregulation can stall glial maturation and promote reactive gliosis or proliferation of undifferentiated glial progenitors. Clinically, elevated phosphorylated MEK/ERK levels in cerebrospinal fluid or biopsy samples could serve as early biomarkers of inflammation-induced gliogenic imbalance. In patients recovering from CNS infections, autoimmune diseases, or brain trauma, monitoring MAPK/MEK pathway activity could help predict long-term neurodevelopmental disruptions or gliosis severity. Diagnostic imaging tools or liquid biopsies paired with molecular profiling may enable early detection of aberrant glial expansion, particularly in pediatric or immunocompromised individuals. Therapeutically, targeting the MAPK/MEK pathway during or after inflammatory episodes may offer a strategy to restore healthy glial differentiation and prevent tumorigenic progression. MEK inhibitors, currently explored in oncology, could be repurposed to mitigate excessive glial progenitor proliferation and promote their proper maturation. In parallel, anti-inflammatory agents that reduce upstream cytokine activity could help normalize pathway signaling. For high-risk populations, such as those with chronic neuroinflammation or prior glial scarring, periodic molecular surveillance of MEK/ERK activation alongside other oncogenic markers could inform preventive care and early therapeutic intervention. The chronic overactivation of this pathway not only fosters an environment conducive to glioma formation by enabling survival of mutation-prone glial progenitors but may also impair the functional roles of mature glia, further compromising brain repair and resilience. Thus, the MAPK/MEK axis represents both a clinical biomarker and a therapeutic target in reducing inflammation-associated glial pathologies and long-term brain tumor risk.

#### 12. E2F/TCFL2 signaling pathway

##### Neuroinflammation-Mediated Dysregulation in Glial Cells

The E2F/TCFL2 signaling pathway plays a pivotal role in regulating the cell cycle, differentiation, and proliferation of glial progenitors during gliogenesis. Under normal conditions, E2F transcription factors, including E2F1, E2F3, and E2F4, control the expression of genes involved in the transition from quiescence to proliferation in glial cells, as well as their subsequent differentiation into mature glial cell types, such as astrocytes and oligodendrocytes. The activity of these transcription factors is tightly regulated by the retinoblastoma protein (Rb) and other cell cycle regulators to ensure that glial progenitors proliferate at appropriate times during development [97]. However, brain inflammation, which is typically characterized by the release of pro-inflammatory cytokines and chemokines, can disrupt this finely balanced regulation, leading to dysregulation of the E2F/TCFL2 pathway. In the context of brain inflammation, the dysregulation of the E2F/TCFL2 signaling pathway can result from the increased release of inflammatory mediators like TNF-α, IL-1β, and IL-6. These cytokines are known to activate various signaling cascades, including NF-κB and MAPK pathways, which can interfere with the normal regulation of the E2F family. For example, inflammatory signaling can lead to the phosphorylation of Rb, which in turn releases E2F transcription factors, allowing them to promote the expression of genes that drive the cell cycle forward. In this environment, glial progenitors may enter an unchecked proliferative state, bypassing critical checkpoints in the differentiation process [98]. The failure to properly regulate E2F activity may impair the differentiation of glial cells, resulting in the accumulation of undifferentiated progenitors. Furthermore, brain inflammation can alter the expression of various cell cycle regulators that interact with E2F/TCFL2 factors. Cyclin-dependent kinases (CDKs), which are crucial for the progression of the cell cycle, can be upregulated during inflammation, leading to excessive phosphorylation of Rb proteins and premature activation of E2F transcription factors. This unchecked activation can lead to excessive proliferation of glial progenitors, preventing them from properly differentiating into mature astrocytes, oligodendrocytes, or other glial cell types. Additionally, inflammation-induced DNA damage and the accumulation of reactive oxygen species (ROS) can further enhance the activation of E2F/TCFL2, compounding the issue by promoting the survival and proliferation of these aberrant glial progenitors. The dysregulation of the E2F/TCFL2 pathway in the setting of brain inflammation may have long-term consequences, including the potential for increased tumorigenesis. Inflammation is often linked to a tumor-promoting microenvironment, where chronic activation of pro-proliferative signaling pathways, such as E2F/TCFL2, creates an environment conducive to the accumulation of genetic mutations and cellular transformation [99].

Prolonged and aberrant activation of E2F may prevent glial progenitors from exiting the cell cycle at appropriate times, allowing them to accumulate genetic errors. These mutations may predispose glial cells to malignant transformation, leading to the formation of gliomas or other types of brain tumors later in life. Additionally, the disruption of normal glial cell differentiation due to the dysregulated E2F/TCFL2 pathway may lead to the persistence of undifferentiated progenitors in the brain. These progenitors are more prone to transformation into tumorigenic cells because they lack the full range of differentiation signals necessary to stabilize their genetic and epigenetic programs. The failure to fully differentiate into mature glial cells may also result in dysfunctional gliogenesis, with abnormal glial populations that are unable to properly support neuronal function and maintain homeostasis in the brain. Such cellular dysfunction may contribute to the development of a more permissive environment for tumorigenesis, particularly in the context of ongoing inflammation and cellular stress. In summary, brain inflammation can lead to the dysregulation of the E2F/TCFL2 pathway, driving glial progenitors into a proliferative state while impairing their differentiation into mature glial cells. This disruption, compounded by the release of inflammatory cytokines and signaling molecules, can lead to excessive cell cycle progression, preventing the proper establishment of glial cell identities. Over time, this dysregulation may result in the accumulation of genetic mutations, increasing the risk of tumorigenesis. The persistence of undifferentiated glial progenitors, along with the unchecked activity of the E2F/TCFL2 pathway, creates a cellular environment that supports the development of gliomas and other brain tumors, highlighting the critical need for controlling inflammation in the brain to maintain proper gliogenesis and prevent tumor formation [100].

##### Consequences for Glial Cell Fate and Long-Term Brain Tumor Risk

Brain inflammation can have a significant impact on the regulation of the E2F/TCFL2 signaling pathway, which plays a key role in the proper development of glial cells during gliogenesis. Under normal conditions, the E2F transcription factors regulate the cell cycle and differentiation of glial progenitor cells by controlling the expression of genes necessary for the transition from proliferation to differentiation. However, inflammation in the brain, characterized by the release of pro-inflammatory cytokines and other immune mediators, can disrupt the delicate balance of this pathway, leading to alterations in glial cell fate. During brain inflammation, cytokines such as IL-1β, TNF-α, and IL-6 can activate various signaling cascades, including NF-κB and MAPK, which in turn can affect the activity of E2F transcription factors [101]. These inflammatory signals often lead to the phosphorylation of retinoblastoma (Rb) proteins, which releases E2F transcription factors from their inhibitory binding and enables them to activate cell cycle-promoting genes. In the context of brain inflammation, this dysregulated activation of E2F can result in excessive proliferation of glial progenitor cells, preventing their differentiation into mature glial cell types such as astrocytes and oligodendrocytes. This interference with differentiation can cause an accumulation of undifferentiated progenitors, which are prone to further genetic alterations. The dysregulation of the E2F/TCFL2 pathway in the presence of inflammation can be further compounded by increased DNA damage, oxidative stress, and reactive oxygen species (ROS) generated in the inflammatory microenvironment. These factors can exacerbate the activation of E2F transcription factors, encouraging the continuation of cell cycle progression without the necessary differentiation cues. When glial progenitors fail to properly exit the cell cycle and differentiate, they remain in a proliferative state, which increases the risk of accumulating mutations over time [102]. This accumulation of genetic errors is a key factor in the development of tumors, as the uncontrolled proliferation of glial progenitors creates an ideal environment for the initiation and progression of tumorigenesis. Furthermore, the persistence of undifferentiated glial progenitors due to the failure of proper E2F-mediated differentiation may contribute to an imbalance in the cellular composition of the brain. This disruption in gliogenesis can result in glial cells that are less capable of supporting neuronal function and maintaining homeostasis. Additionally, the continuous activation of the E2F pathway due to prolonged inflammation may increase the likelihood of glial progenitors becoming tumorigenic. The abnormal accumulation of these progenitors, coupled with ongoing inflammation and genetic instability, creates a microenvironment conducive to the development of brain tumors, particularly gliomas [103]. Brain inflammation can significantly disrupt the E2F/TCFL2 signaling pathway, leading to the unchecked proliferation of glial progenitors and their failure to differentiate into mature glial cells. This dysregulation, driven by inflammatory cytokines and cellular stressors, can result in an accumulation of genetic mutations and an increased risk of brain tumor formation [104].

##### Possible Clinical Implications in Patients: Toward Improved Diagnosis, Surveillance, and Therapeutics

Normally responsible for orchestrating the cell cycle and ensuring proper glial progenitor proliferation and differentiation, the E2F pathway becomes destabilized by inflammatory mediators like IL-1β, IL-6, and TNF-α. This leads to unchecked glial proliferation and impaired differentiation, a hallmark of gliogenic imbalance. In clinical settings, persistent upregulation of E2F target genes or hyperphosphorylation of the Rb protein may serve as biomarkers for glial dysregulation in patients with chronic CNS inflammation. Such markers could be detected via cerebrospinal fluid (CSF) analysis or molecular imaging, providing early warning signs of aberrant gliogenesis in individuals recovering from encephalitis, neurodegenerative disorders, or traumatic brain injury. By integrating E2F activity profiling into post-inflammatory surveillance programs, clinicians could better predict which patients are at elevated risk for tumor-prone glial behavior or delayed neurological recovery. From a therapeutic perspective, modulating E2F/TCFL2 pathway activity presents a promising strategy to prevent the long-term consequences of neuroinflammation, including gliomagenesis. Agents that restore Rb-E2F checkpoint function or inhibit downstream pro-proliferative targets may help re-establish the balance between glial proliferation and differentiation.

Additionally, anti-inflammatory interventions aimed at reducing NF-κB and MAPK signaling could indirectly stabilize E2F activity by minimizing cytokine-driven Rb phosphorylation. For high-risk patients, especially those with persistent inflammatory markers and abnormal glial proliferation, regular surveillance using transcriptomic or epigenetic assays of E2F and its regulatory network could enable early intervention before tumorigenic transformations occur.

Moreover, therapies targeting oxidative stress and DNA repair pathways may be critical for reducing the mutational burden in glial progenitor pools exposed to chronic inflammation. Overall, understanding how the E2F/TCFL2 axis is altered in neuroinflammatory conditions opens avenues for early diagnosis, precise risk stratification, and novel treatments that can reduce the incidence of inflammation-induced brain tumors such as gliomas.

#### 13. NFIX/Ephrins/Netrins signaling pathway Neuroinflammation-Mediated Dysregulation in Glial Cells

The NFIX/Ephrins/Netrins signaling pathway plays an essential role in the developmental biology of gliogenesis by influencing the migration, differentiation, and survival of glial cells, particularly during early brain development. NFIX, a transcription factor, interacts with ephrins and netrins, which are guidance cues that help in the proper spatial arrangement and differentiation of glial cells. Ephrins and netrins are involved in establishing cell-cell communication and guiding cell migration, while NFIX acts as a key regulator of glial progenitor differentiation and maturation.

However, during brain inflammation, this intricate network of signals can become dysregulated due to the heightened activity of pro-inflammatory cytokines and immune mediators, leading to significant disruptions in glial cell fate and gliogenesis [105]. Brain inflammation is typically characterized by the activation of microglia, the brain’s resident immune cells, as well as the release of inflammatory cytokines such as TNF-α, IL-1β, and IL-6. These inflammatory mediators can alter the expression and function of various signaling molecules involved in gliogenesis, including NFIX, ephrins, and netrins. For example, prolonged exposure to inflammatory signals can lead to changes in the expression of NFIX, potentially impairing its regulatory role in glial progenitor differentiation. Inflammatory cytokines can interfere with the ability of NFIX to activate the transcription of genes required for glial cell differentiation, thereby preventing glial progenitors from progressing toward their intended cell fates, such as becoming mature astrocytes or oligodendrocytes. Similarly, the signaling pathways involving ephrins and netrins, which are crucial for guiding glial cells to their appropriate locations and ensuring proper differentiation, can be significantly altered during brain inflammation. Inflammatory cytokines have been shown to impact the expression of ephrin receptors and netrin receptors on glial progenitor cells, disrupting their ability to respond to these essential guidance cues [106]. This disruption can lead to improper cell migration, aberrant glial cell positioning, and failure of differentiation, as glial progenitors are unable to migrate to the correct locations or respond to the necessary differentiation signals.

Additionally, brain inflammation may result in the increased activation of reactive oxygen species (ROS) and other stressors in the brain, further exacerbating the dysregulation of NFIX and its associated signaling pathways. Oxidative stress can lead to the alteration of protein function and cellular signaling, impairing the ability of NFIX to regulate glial progenitor fate. Moreover, the persistent inflammatory environment may cause prolonged activation of pro-inflammatory pathways such as NF-κB, which can further interfere with the expression and function of NFIX, ephrins, and netrins, exacerbating the dysfunction in gliogenesis. The dysregulation of the NFIX/Ephrins/Netrins signaling pathway due to brain inflammation has significant implications for glial cell development and function. If glial progenitors fail to properly differentiate or migrate due to the disruption of these pathways, it can result in an accumulation of undifferentiated or malfunctioning glial cells. These cells may harbor genetic mutations or undergo abnormal proliferative processes, creating a permissive environment for the development of gliomas or other brain tumors [107]. The persistence of abnormal glial progenitors that are unable to differentiate into functional glial cells could also contribute to neurodegenerative conditions, further compounding the negative effects of brain inflammation on brain health. In conclusion, brain inflammation can disrupt the NFIX/Ephrins/Netrins signaling pathway by altering the expression and function of NFIX and its associated guidance cues. These disruptions can impair the differentiation, migration, and survival of glial cells, increasing the risk of gliomas and other brain tumors.

The dysregulation of this pathway, driven by inflammatory cytokines, oxidative stress, and immune activation, may not only interfere with normal gliogenesis but also contribute to the long-term pathology of the brain by promoting abnormal glial cell behavior, genetic instability, and tumorigenesis [108].

##### Consequences for Glial Cell Fate and Long-Term Brain Tumor Risk

Brain inflammation can significantly impact the NFIX/Ephrins/Netrins signaling pathways, disrupting the proper cell fate determination of glial cells and potentially increasing the risk of brain tumors later in life. The NFIX gene, which plays a crucial role in regulating the differentiation of glial progenitors, acts through interactions with signaling cues such as ephrins and netrins, which guide the migration and fate decisions of these cells. Ephrins, through their Eph receptor interactions, and netrins, by binding to their corresponding receptors, influence various cellular processes, including migration, survival, and differentiation [109]. During inflammation, the upregulation of pro-inflammatory cytokines, such as TNF-α, IL-1β, and IL-6, as well as the activation of microglia, can profoundly affect the activity and expression of these signaling molecules, leading to impaired glial development. In an inflammatory environment, the excessive production of inflammatory mediators can lead to alterations in the expression of NFIX, which in turn may affect the differentiation of glial progenitors. NFIX is a key transcription factor that drives the maturation of glial cells such as astrocytes and oligodendrocytes. Inflammatory signals can disrupt NFIX expression or its ability to activate critical differentiation pathways, leaving glial progenitors in an undifferentiated or dysfunctional state [110]. This dysregulation may lead to impaired maturation and function of glial cells, preventing them from performing their roles in maintaining neural homeostasis and promoting normal brain function. Furthermore, disruptions in NFIX activity may prevent glial progenitors from responding properly to the guidance cues provided by ephrins and netrins, affecting their migration to appropriate locations within the brain and impairing their differentiation into mature glial subtypes. The ephrins and netrins themselves can also be dysregulated in the context of brain inflammation.

Inflammatory cytokines can alter the expression of ephrin and netrin receptors on glial progenitors, which may disrupt their ability to correctly interpret and respond to these guidance cues. Ephrin and netrin signaling are crucial for ensuring that glial progenitors migrate to their proper locations and differentiate into specific glial cell types. The disruption of this guidance system could result in improper migration, abnormal positioning, or a failure of glial progenitors to mature into functional astrocytes or oligodendrocytes. Such misdirected migration or differentiation can contribute to the development of gliomas or other types of brain tumors, as undifferentiated or poorly differentiated cells may acquire proliferative capabilities or genetic mutations that drive tumorigenesis. In addition to these effects on cell fate determination, brain inflammation can promote an environment of increased oxidative stress and the accumulation of reactive oxygen species (ROS). This stress may further interfere with the proper signaling of NFIX, ephrins, and netrins, impairing the capacity of glial progenitors to undergo normal differentiation. The chronic activation of pro-inflammatory signaling pathways, such as NF-κB, in response to brain inflammation can exacerbate this dysregulation, resulting in prolonged disruption of glial cell maturation and an increased likelihood of cellular mutations [111]. These mutations, if occurring in key regulatory genes involved in glial differentiation or cell cycle control, could increase the risk of brain tumor formation, particularly gliomas, later in life. Moreover, the persistence of abnormal glial progenitors, which are unable to fully differentiate into mature glial cells, can create a pool of cells that exhibit genetic instability. These cells may evade normal differentiation signals and adopt a more proliferative phenotype, increasing the likelihood of tumorigenesis. Inflammatory-induced changes in the NFIX/Ephrins/Netrins pathway can create an environment conducive to this type of cellular behavior, where a failure in cell fate decisions or response to guidance cues allows cells to proliferate uncontrollably. Over time, such cellular dysfunctions could accumulate, leading to the emergence of gliomas or other brain tumors. In conclusion, brain inflammation can significantly disrupt the NFIX/Ephrins/Netrins signaling pathways by altering the expression and function of key regulatory genes and guidance molecules involved in gliogenesis. This disruption impairs the ability of glial progenitors to migrate, differentiate, and mature properly, resulting in an accumulation of undifferentiated or dysfunctional glial cells. These abnormalities, compounded by the effects of oxidative stress and chronic inflammatory signaling, may increase the risk of brain tumors, particularly gliomas, later in life. The dysregulation of NFIX, ephrins, and netrins in the context of brain inflammation provides a pathway through which glial cell fate decisions are disrupted, and cellular instability is promoted, thereby contributing to the potential for tumorigenesis in the brain [112].

##### Possible Clinical Implications in Patients: Toward Improved Diagnosis, Surveillance, and Therapeutics

The dysregulation of the NFIX/Ephrins/Netrins signaling pathway in response to brain inflammation holds important clinical implications for early diagnosis, therapeutic targeting, and long-term surveillance in neuroinflammatory and neuro-oncologic settings. Under normal physiological conditions, this pathway orchestrates critical aspects of gliogenesis, including the spatial migration, differentiation, and maturation of glial progenitor cells. However, in inflamed neural environments, elevated cytokines such as IL-1β and TNF-α can downregulate NFIX expression and disrupt ephrin/netrin signaling, impeding glial migration and proper differentiation. Clinically, this disruption may be reflected in developmental delays, demyelinating conditions, or early gliotic responses observable via advanced neuroimaging or cerebrospinal fluid biomarkers. Measuring NFIX levels or altered ephrin/netrin receptor profiles could thus offer a valuable diagnostic window into early-stage glial mispatterning or inflammatory injury in the developing or adult brain. In pediatric patients with encephalitis or in individuals with chronic neuroinflammation, screening for dysregulation of this axis may help identify those at elevated risk of delayed recovery or progression to gliosis or glioma. Therapeutically, the NFIX/Ephrins/Netrins axis represents a promising target for intervention aimed at restoring healthy glial development and preventing long-term complications. Pharmacological agents that modulate ephrin or netrin receptor signaling or gene therapy approaches to normalize NFIX expression could be explored to reestablish glial migratory and differentiation dynamics following inflammation. In neuro-oncology, long-term surveillance of patients with chronic CNS inflammation could include molecular monitoring of NFIX and associated guidance molecules as part of a predictive model for glioma risk. Since undifferentiated glial progenitors disrupted by this pathway are more likely to accumulate mutations and resist apoptotic signals, therapies that promote terminal differentiation or reduce oxidative stress may also help prevent malignant transformation.

Ultimately, integrating this molecular pathway into clinical practice could aid in developing precision-based strategies for early intervention, particularly in high-risk neuroinflammatory conditions that predispose to glial malignancies.

## Discussion

### Possible Contribution Toward Advances in Future Patient Care Through This Study

1. **Early Diagnosis and Risk Stratification**

a. Identification of key gliogenic pathways disrupted by neuroinflammation (e.g., FGFR3, STAT3, IL-6, NF-κB) may enable the development of early diagnostic biomarkers.
b. Patients recovering from CNS infections (e.g., meningitis, encephalitis) or suffering from chronic neuroinflammatory disorders could be monitored for glial pathway abnormalities to assess future neurological or oncological risks.
c. Molecular screening tools targeting these pathways could help stratify patients based on susceptibility to gliosis, neurodevelopmental delay, or glial-derived tumors.
2. Targeted Therapeutic Development

This study highlights therapeutic targets within glial signaling cascades, such as JAK-STAT inhibition, FGFR3 modulation, and NF-κB pathway stabilization.
Restoring normal glial differentiation and suppressing aberrant progenitor proliferation through these targets could mitigate long-term neuroinflammatory damage and reduce glioma risk.
Future pharmacological agents may be designed to counteract inflammation-induced pathway dysregulation in high-risk individuals.
3. Personalized Monitoring and Surveillance

Patients with a history of neuroinflammation could benefit from longitudinal molecular surveillance of glial regulatory genes and proteins.
Monitoring for persistent activation of tumorigenic pathways (e.g., STAT3, E2F, MEK/ERK) may allow clinicians to detect early precancerous changes in glial populations.
Surveillance protocols based on pathway profiling could be integrated into care plans for immunocompromised or pediatric populations vulnerable to CNS inflammation.
4. Neuroprotective and Regenerative Strategies

Understanding how inflammation interferes with gliogenesis opens avenues for therapies that promote repair, remyelination, and functional glial recovery post-injury.
Interventions aimed at modulating ephrin/netrin or Neuregulin-1 signaling could enhance neural-glial communication and tissue regeneration.
Such therapies could improve cognitive and motor outcomes in patients with inflammatory CNS injuries or post-infectious sequelae.
5. Prevention of Glial Tumorigenesis

By linking glial fate dysregulation to tumorigenic transformation, the study supports preventative strategies in neuro-oncology.
Therapeutic modulation of glial signaling pathways may reduce the accumulation of undifferentiated, mutation-prone glial progenitors.
. Ultimately, this knowledge can contribute to lowering the lifetime risk of developing gliomas in inflammation-prone individuals.

### Long-Term Impact of Early-Life Glial Inflammation on Brainstem Cardio-Pulmonary Centers

Early-life glial inflammation can exert lasting effects on brainstem regions that regulate autonomic functions, particularly the medulla oblongata, which houses critical cardio-pulmonary centers such as the nucleus tractus solitarius (NTS) and ventrolateral medulla (VLM). During neurodevelopment, glial cells especially astrocytes and microglia play essential roles in shaping synaptic connectivity and homeostatic regulation within these regions. However, if inflammation occurs during critical developmental windows, cytokine-mediated dysregulation of gliogenesis can alter the maturation and function of these glial networks. This can impair astrocyte-neuron interactions essential for baroreflex sensitivity, chemoreceptor responsiveness, and the maintenance of respiratory rhythm. Sustained activation of glial cells due to early neuroinflammation may also promote long-term neuroimmune priming, oxidative stress, and aberrant glial remodeling in autonomic circuits.

Over time, these changes may predispose individuals to impaired cardio-respiratory control, increasing susceptibility to hypertension, arrhythmias, and sudden cardiac or respiratory events, especially under stress or hypoxic conditions. This suggests that inflammation-induced glial dysregulation in early life is not only a neurodevelopmental concern but also a potential driver of cardiovascular and pulmonary morbidity in adulthood, underscoring the need for surveillance and early interventions in at-risk populations.

### Clinical Implications and Implications Towards Future Research Directions

The findings of this research highlight significant clinical implications for understanding how brain inflammation affects gliogenic regulators and their role in glial cell fate decisions. By identifying how inflammation disrupts key signaling pathways, such as FGFR3, JAK-STAT, STAT3, and others, this research can inform potential therapeutic strategies aimed at modulating these pathways to prevent or treat glioma and other brain tumors. Targeting the molecular players involved in inflammation-driven gliogenesis may offer novel approaches to slowing tumor progression or even preventing tumor initiation in high-risk populations. Moreover, this research opens the door to precision medicine by enabling the development of biomarkers for early detection of abnormal glial differentiation due to inflammation. Detecting disruptions in signaling pathways like NF-κB and IL-6 could allow clinicians to identify patients at higher risk for developing brain tumors, facilitating earlier interventions and improved outcomes. The identification of inflammation-induced changes in gliogenesis also underscores the need for further research into the timing, intensity, and chronicity of brain inflammation that leads to dysregulated glial differentiation and tumor formation. Future research should aim to elucidate the molecular mechanisms driving the chronic activation of these signaling pathways during inflammation and explore potential therapeutic inhibitors or modulators to correct the disrupted signaling in glial cells. Further studies could also investigate the role of the brain’s immune microenvironment, which may play a central role in the development of tumor-promoting inflammation. Understanding how specific inflammatory cytokines or environmental factors affect gliogenic pathways will be essential for designing more targeted interventions. In addition, research should focus on the long-term effects of inflammation-induced gliogenesis across different age groups and its association with neurodegenerative diseases. Investigating the impact of inflammation on glial cell aging and how it relates to glioma risk in the elderly population could provide insights into the role of chronic neuroinflammation in age-related brain disorders. Expanding the scope of this research into pre-clinical models and clinical trials will also be crucial in translating these findings into effective therapies and preventative measures for brain tumor development.

### Inflammation disrupting Proliferation Vs Differentiation Balance

Brain inflammation disrupts the balance between glial cell proliferation and differentiation by altering key gliogenic regulators. Inflammatory mediators such as cytokines and reactive molecules can directly affect pathways that control both cell proliferation and differentiation, resulting in an environment where glial cells are either unable to differentiate properly or continue proliferating uncontrollably. For instance, signaling through FGFR3 can promote the proliferation of glial progenitor cells, but under inflammatory conditions, this pathway may become dysregulated, skewing the balance toward unchecked proliferation rather than the appropriate differentiation of glial cells. Similarly, the JAK-STAT pathway, particularly the activation of STAT3, can be influenced by inflammation, leading to the promotion of glial cell proliferation rather than differentiation. This occurs because STAT3 activation is often linked to a proliferative response, and in the context of inflammation, this response can be exaggerated, impairing the differentiation potential of glial progenitors and promoting their expansion. S100, a calcium-binding protein, plays a role in the regulation of glial differentiation, but under inflammatory conditions, its expression can be altered, leading to the maintenance of progenitor cell states and hindered differentiation.

Hey1, a key regulator of the Notch signaling pathway, also becomes impacted by inflammation, which can result in the inhibition of glial cell differentiation. Notch signaling, often active during the proliferation phase of gliogenesis, may persist inappropriately under inflammatory stress, thus delaying differentiation and continuing the cell cycle. Likewise, the HES1 transcription factor, which is downstream of Notch signaling, may be upregulated by inflammation, promoting a proliferative, undifferentiated state in glial progenitors. This disturbance creates a feedback loop where the lack of differentiation further supports the proliferative environment, increasing the number of undifferentiated glial cells. Inflammation also disrupts the function of DTX, a regulator of Notch signaling, which normally helps coordinate the transition from proliferation to differentiation. When this pathway is dysregulated during inflammation, the differentiation process can be delayed or blocked altogether, increasing the pool of proliferating progenitors. Similarly, inflammatory cytokines like IL-6 can activate NF-κB, a central mediator of inflammatory responses. The activation of NF-κB has been shown to promote cell cycle progression and inhibit the differentiation of glial cells. Inflammatory signaling through NF-κB can thus tip the balance toward proliferation by blocking differentiation and favoring the maintenance of a proliferative state. Neuregulin-1, involved in the development of glial cells, can also be impacted by inflammation, altering signaling that regulates both proliferation and differentiation, resulting in an imbalance that favors the expansion of progenitor cells at the expense of their maturation into differentiated glial cells. Additionally, the MAPK-MEK pathway, which is involved in cellular responses to growth factors and stress signals, becomes hyperactivated during brain inflammation. This activation promotes glial progenitor proliferation, while suppressing differentiation signals, leading to an accumulation of undifferentiated progenitors. The E2F/TCFL2 pathway, which regulates the cell cycle, can also be disrupted by inflammation, leading to unchecked cell division. Inflammatory cytokines can affect the activity of these transcription factors, causing a shift from a differentiation-promoting environment to one that favors continuous cell division. Finally, NFIX, Ephrins, and Netrins, important regulators of cell migration and differentiation, are also susceptible to inflammatory disruption. Inflammatory signals may alter the expression of these regulators, impeding proper glial cell migration and differentiation and potentially contributing to an expansion of proliferating progenitors rather than differentiated glial populations. In summary, brain inflammation induces a complex series of dysregulations across multiple gliogenic pathways, including FGFR3, JAK-STAT, STAT3, S100, Hey1, HES1, DTX, IL-6, NF-κB, Neuregulin-1, MAPK-MEK, E2F/TCFL2, and NFIX/Ephrins/Netrins. These disruptions ultimately shift the balance in favor of glial cell proliferation over differentiation, resulting in the expansion of progenitor cells and possibly creating an environment conducive to tumor formation. This imbalance not only affects normal gliogenesis but also contributes to the increased risk of brain tumors, particularly gliomas, by maintaining an undifferentiated, proliferative state in glial cells.

### Conclusion

Inflammation within the brain, driven by pro-inflammatory cytokines and microglial activation, can cause dysregulation in the expression and function of critical signaling pathways and transcription factors involved in gliogenesis. These include FGFR3, JAK-STAT, STAT3, S100, Hey1, HES1, DTX, IL-6, NF-κB, Neuregulin-1, MAPK-MEK, E2F-TCFL2, and NFIX/Ephrins/Netrins, all of which are essential for proper glial cell differentiation, migration, and maturation. The disruption of FGFR3 signaling, for example, impairs the differentiation of glial progenitors, potentially leaving them in a proliferative or undifferentiated state, which may increase susceptibility to tumorigenesis. Similarly, the JAK-STAT and STAT3 pathways, which are vital for regulating the proliferation and differentiation of glial cells, can be altered by inflammatory cytokines, leading to improper glial cell fate decisions and abnormal cell growth. The dysregulation of S100 proteins and other glial-specific markers further contributes to abnormal glial differentiation and maturation, while factors like Hey1 and HES1, which regulate cell fate through Notch signaling, can be misregulated by inflammation, disrupting the balance between glial progenitor expansion and differentiation. Moreover, inflammatory signaling pathways such as IL-6, NF-κB, and MAPK-MEK are known to promote cellular stress and interfere with the normal function of glial progenitors. The chronic activation of NF-κB, in particular, has been implicated in promoting an environment of genetic instability and altered cell cycle regulation, which increases the likelihood of tumorigenesis. Inflammation-driven activation of these pathways can lead to the accumulation of reactive oxygen species (ROS) and oxidative stress, further exacerbating the risk of genetic mutations in glial progenitors. This results in the expansion of abnormal glial cells, which may acquire proliferative properties, thereby increasing the risk of developing brain tumors such as gliomas later in life. Similarly, the disruption of signaling molecules like Neuregulin-1, MAPK-MEK, and E2F-TCFL2 can impair glial progenitor differentiation and migration, leading to aberrant glial cell formation. This can create an unstable environment where glial progenitors remain in an undifferentiated or dysfunctional state, contributing to cellular transformation and tumor formation. Additionally, alterations in the NFIX/Ephrins/Netrins pathway, which regulate the migration and differentiation of glial cells, may exacerbate these issues, causing glial progenitors to fail in their migration to the appropriate brain regions and preventing their proper differentiation into mature glial cell types. Brain inflammation profoundly impacts gliogenic regulators by disrupting the delicate balance required for glial cell differentiation, migration, and maturation. The resulting dysregulation of key signaling pathways and transcription factors creates an environment conducive to abnormal glial cell behavior, including impaired differentiation, cell fate misguidance, and increased proliferation. These disturbances not only affect the proper development and function of glial cells but also heighten the risk of gliomas and other brain tumors later in life. The combination of altered signaling, genetic instability, and cellular dysfunction caused by brain inflammation points to the critical need for understanding the long-term consequences of inflammation in the brain, particularly in the context of glial inflammation and tumorigenesis to improve future patient outcomes.

## Declarations

### Ethics declarations

**Ethics approval and consent to participate**

Not applicable.

### Consent for publication

Not applicable.

### Data Availability statement

All data generated or analyzed during this study are included in this article.

## Acknowledgements

We are thankful to Muhammad Danial Yaqub (MDY), MBBS, for coordinating this research project among multiple co-authors.

## Authors’ Information

**1. Ovais Shafi (OS)\*** is the author of the study and was involved in the idea, concept, design, and methodology of the study, literature search and references. He did the writing, editing, and revision of the manuscript. He was involved in drawing the findings, results, conclusions, implications of the study, interpretation of the data and was involved in all aspects of the study. He prepared and wrote discussion, results, conclusions and all areas of the study. OS extracted and analyzed the data. He was involved in critical evaluation, audit of every aspect of the study, data extraction, adherence of the study to relevant PRISMA guidelines, limitations of the study, references, and all others. He was involved in drawing PRISMA Flow Diagram. The author read and approved the manuscript.

He investigated how the brain inflammation induced disruption of gliogenic regulators (FGFR3, JAK-STAT, STAT3, S100, Hey1, HES1, DTX, IL-6, NF-κB, Neuregulin-1, MAPK/MEK, E2F/TCFL2, NFIX/Ephrins/Netrins) resulting in glial cell fate dysregulations and its contribution to brain tumor risk. He contributed towards all the 100% of investigated areas.

**Ovais Shafi (OS)\***, MBBS - Sindh Medical College - Dow University of Health Sciences, Karachi, Pakistan. He aspires to become an eminent ‘Physician Scientist’. He is devoted to the research in disease development mechanisms, disease origins and therapeutics. OS is also passionate about multiple research areas including clinical trials, clinical medicine, therapeutics, regenerative medicine, precision medicine including gene therapies, finding disease specific targets for gene therapy, role of disease genomics and epigenetics in diagnosis, management, and therapeutics development. He is dedicated to the field of research and clinical medicine.

**Corresponding author: OS**

Correspondence to <u>Ovais Shafi</u>

**2. Muhammad Waqas (MW)** is the co-author of the study. He contributed to the results and conclusions of the study, also contributed to the writing and editing of these sections. He also contributed to the references. He contributed to investigating how the brain inflammation induced disruption of gliogenic regulators (FGFR3, JAK-STAT, STAT3, S100, Hey1, HES1, DTX, IL-6, NF-κB, Neuregulin-1, MAPK/MEK, E2F/TCFL2, NFIX/Ephrins/Netrins) resulting in glial cell fate dysregulations and its contribution to brain tumor risk. He contributed towards all the 100% of investigated areas.

Muhammad Waqas, MBBS - Sindh Medical College - Dow University of Health Sciences, Karachi, Pakistan. He is ECFMG Certified. His future goals include residency in Internal Medicine/Neurology, and fellowship in Cardiology/Nephrology/Critical Care.

**3. Luqman Naseer Virk (LNV)** is the co-author of the study. He contributed to the results and conclusions of the study, also contributed to the writing and editing of these sections. He also contributed to the references. He contributed to investigating how the brain inflammation induced disruption of gliogenic regulators (FGFR3, JAK-STAT, STAT3, S100, Hey1, HES1, DTX, IL-6, NF-κB, Neuregulin-1, MAPK/MEK, E2F/TCFL2, NFIX/Ephrins/Netrins) resulting in glial cell fate dysregulations and its contribution to brain tumor risk. He contributed towards all the 100% of investigated areas.

Luqman Naseer Virk (LNV), MBBS - Sindh Medical College - Dow University of Health Sciences, Karachi, Pakistan. He is passionate doctor and is enthusiastic about research in medicine/surgery and disease development mechanisms including gastroenterology, cardiology, neurodegenerative diseases, oncogenesis and others.

**4. Sameed Abdul Hameed Siddiqui (SAHS)** is the co-author of the study. He contributed to the results and conclusions of the study, also contributed to the writing and editing of these sections. He also contributed to the references. He contributed to investigating how the brain inflammation induced disruption of gliogenic regulators (FGFR3, JAK-STAT, STAT3, S100, Hey1, HES1, DTX, IL-6, NF-κB, Neuregulin-1, MAPK/MEK, E2F/TCFL2, NFIX/Ephrins/Netrins) resulting in glial cell fate dysregulations and its contribution to brain tumor risk. He contributed towards all the 100% of investigated areas. He was also involved in the audit and critical evaluation of the study.

Sameed Abdul Hameed Siddiqui, Internal Medicine Trainee, Lincoln County Hospital, United Lincolnshire Hospitals NHS Trust, UK.

**5. Osama Jawed Khan (OJK)** is the co-author of the study. He contributed to the results and conclusions of the study, also contributed to the writing and editing of these sections. He also contributed to the references. He contributed to investigating how the brain inflammation induced disruption of gliogenic regulators (FGFR3, JAK-STAT, STAT3, S100, Hey1, HES1, DTX, IL-6, NF-κB, Neuregulin-1, MAPK/MEK, E2F/TCFL2, NFIX/Ephrins/Netrins) resulting in glial cell fate dysregulations and its contribution to brain tumor risk. He contributed towards all the 100% of investigated areas.

Osama Jawed Khan, MBBS - Sindh Medical College - Dow University of Health Sciences, Karachi, Pakistan. He has been affiliated with Dr. Ruth K. M. Pfau, Civil Hospital Karachi, Pakistan. Currently, he is affiliated with Royal Institute of Medicine & Surgery (RIMS), Karachi. He intends to pursue fellowship of the Royal Colleges of Surgeons (FRCS).

**6. Ibrahim Abdul Rahman (IAR)** is the co-author of the study. He contributed to the results and conclusions of the study, also contributed to the writing and editing of these sections. He also contributed to the references. He contributed to investigating how the brain inflammation induced disruption of gliogenic regulators (FGFR3, JAK-STAT, STAT3, S100, Hey1, HES1, DTX, IL-6, NF-κB, Neuregulin-1, MAPK/MEK, E2F/TCFL2, NFIX/Ephrins/Netrins) resulting in glial cell fate dysregulations and its contribution to brain tumor risk. He contributed towards all the 100% of investigated areas.

He is a passionate doctor and is enthusiastic about research in neurodegenerative diseases, psychiatry, cognition and other areas. His goal is to make significant impact in the field of Research.

**7. Ameerah Karim (AK)** is the co-author of the study. She contributed to the results and conclusions of the study, also contributed to the writing and editing of these sections. She also contributed to the references. She contributed to investigating how the brain inflammation induced disruption of gliogenic regulators (FGFR3, JAK-STAT, STAT3, S100, Hey1, HES1, DTX, IL-6, NF-κB, Neuregulin-1, MAPK/MEK, E2F/TCFL2, NFIX/Ephrins/Netrins) resulting in glial cell fate dysregulations and its contribution to brain tumor risk. She contributed towards all the 100% of investigated areas.

Ameerah Karim - Lincoln County Hospital, Lincoln, United Kingdom

**8. Muhammad Danyal Khalid (MDK)** is the co-author of the study. He contributed to the results and conclusions of the study, also contributed to the writing and editing of these sections. He also contributed to the references. He contributed to investigating how the brain inflammation induced disruption of gliogenic regulators (FGFR3, JAK-STAT, STAT3, S100, Hey1, HES1, DTX, IL-6, NF-κB, Neuregulin-1, MAPK/MEK, E2F/TCFL2, NFIX/Ephrins/Netrins) resulting in glial cell fate dysregulations and its contribution to brain tumor risk. He contributed towards all the 100% of investigated areas.

He is currently working at Avicenna Medical College and graduated from Lahore Medical and Dental College, Pakistan.

**9. Maleeha Anum (MA)** is the co-author of the study. She contributed to the results and conclusions of the study, also contributed to the writing and editing of these sections. She also contributed to the references. She contributed to investigating how the brain inflammation induced disruption of gliogenic regulators (FGFR3, JAK-STAT, STAT3, S100, Hey1, HES1, DTX, IL-6, NF-κB, NFIX/Ephrins/Netrins) resulting in glial cell fate dysregulations and its contribution to brain tumor risk. She contributed towards the 80% of investigated area.

Maleeha Anum, MBBS, Jinnah Sindh Medical University, Karachi, Pakistan.

**10. Sameeta Makwana (SM)** is the co-author of the study. She contributed to the results and conclusions of the study, also contributed to the writing and editing of these sections. She also contributed to the references. She contributed to investigating how the brain inflammation induced disruption of gliogenic regulators (FGFR3, JAK-STAT, STAT3, S100, Hey1, HES1, DTX, IL-6, NF-κB, NFIX/Ephrins/Netrins) resulting in glial cell fate dysregulations and Its contribution to brain tumor risk. She contributed towards the 80% of investigated areas.

Sameeta Makwana, MBBS, Jinnah Sindh Medical University, Karachi, Pakistan.

**11. Manesh Kumar (MK)** is the co-author of the study. He contributed to the results and conclusions of the study, also contributed to the writing and editing of these sections. He also contributed to the references. He contributed to investigating how the brain inflammation induced disruption of gliogenic regulators (FGFR3, JAK-STAT, STAT3, IL-6, NF-κB, Neuregulin-1, MAPK/MEK, E2F/TCFL2, NFIX/Ephrins/Netrins) resulting in glial cell fate dysregulations and Its contribution to brain tumor risk. He contributed towards the 70% of investigated areas.

Manesh Kumar, MBBS, Shaheed Mohtarma Benazir Bhutto Medical University, Larkana, Pakistan

**12. Aamir Ameer (AA)** is the co-author of the study. He contributed to the results and conclusions of the study, also contributed to the writing and editing of these sections. He also contributed to the references. He contributed to investigating how the brain inflammation induced disruption of gliogenic regulators (FGFR3, JAK-STAT, STAT3, IL-6, NF-κB, Neuregulin-1, MAPK/MEK, E2F/TCFL2, NFIX/Ephrins/Netrins) resulting in glial cell fate dysregulations and Its contribution to brain tumor risk. He contributed towards the 70% of investigated areas.

Aamir Ameer, MBBS, Jinnah Sindh Medical University, Karachi, Pakistan.

**13. Finza Kanwal (FK)** is the co-author of the study. She contributed to the results and conclusions of the study, also contributed to the writing and editing of these sections. She also contributed to the references. She contributed to investigating how the brain inflammation induced disruption of gliogenic regulators (FGFR3, JAK-STAT, STAT3, S100, Hey1, HES1, DTX, IL-6, NF-κB, Neuregulin-1, MAPK/MEK, E2F/TCFL2, NFIX/Ephrins/Netrins) resulting in glial cell fate dysregulations and Its contribution to brain tumor risk. She contributed towards all the 100% of investigated areas.

Finza Kanwal, MBBS - Sindh Medical College - Dow University of Health Sciences, Karachi, Pakistan. She has been affiliated with Liaquat National Hospital and Medical College, Pakistan. Currently, she is affiliated with The College of Physicians and Surgeons Pakistan. Her goal is to be an Endocrinologist (MRCP).

**14. Raveena (RA)** is the co-author of the study. She contributed to the results and conclusions of the study, also contributed to the writing and editing of these sections. She also contributed to the references. She contributed to investigating how the brain inflammation induced disruption of gliogenic regulators (FGFR3, JAK-STAT, STAT3, S100, Hey1, HES1, DTX, IL-6, NF-κB, Neuregulin-1, MAPK/MEK, E2F/TCFL2, NFIX/Ephrins/Netrins) resulting in glial cell fate dysregulations and Its contribution to brain tumor risk. She contributed towards all the 100% of investigated areas.

Raveena, MBBS - Sindh Medical College – Jinnah Sindh Medical University, Karachi, Pakistan. She is passionate about research in surgery and disease development mechanisms including neurodegenerative diseases, oncogenesis and others. She is ECFMG Certified. She is passionate about residency in Internal Medicine/Surgery. Her goal is to make significant impact in the field of Research.

**15. Javeryah Rafiq Shaikh (JRS)** is the co-author of the study. She contributed to the results and conclusions of the study, also contributed to the writing and editing of these sections. She contributed to investigating how the brain inflammation induced disruption of gliogenic regulators (FGFR3, JAK-STAT, STAT3, S100, HES1, DTX, E2F/TCFL2, NFIX/Ephrins/Netrins) resulting in glial cell fate dysregulations and Its contribution to brain tumor risk. She contributed towards the 77% of investigated areas. Javeryah Rafiq Shaikh, MBBS - Jinnah Sindh Medical University, Karachi, Pakistan.
**16. Muhammad Danial Yaqub (MDY)** is the co-author of the study. He contributed to the results and conclusions of the study, also contributed to the writing and editing of these sections. He also contributed to the references. He contributed to investigating how the brain inflammation induced disruption of gliogenic regulators (FGFR3, JAK-STAT, STAT3, S100, Hey1, HES1, DTX, IL-6, NF-κB, Neuregulin-1, MAPK/MEK, E2F/TCFL2, NFIX/Ephrins/Netrins) resulting in glial cell fate dysregulations and Its contribution to brain tumor risk. He contributed towards all the 100% of investigated areas.

Muhammad Danial Yaqub, MBBS, Lincoln County Hospital, Lincoln, United Kingdom. He has keen interests in Clinical Medicine. MDY aspires to be a researcher which can present new data which can pave the way for early diagnosis and treatment. His goal is also to translate emerging findings in research of disease development mechanisms/ origins into their clinical implications.

**17. Ayesha Saeed (AS)** is the co-author of the study. She contributed to the results and conclusions of the study, also contributed to the writing and editing of these sections. She also contributed to the references. She contributed to investigating how the brain inflammation induced disruption of gliogenic regulators (FGFR3, JAK-STAT, STAT3, S100, Hey1, HES1, DTX, IL-6, NF-κB, Neuregulin-1, MAPK/MEK, E2F/TCFL2, NFIX/Ephrins/Netrins) resulting in glial cell fate dysregulations and Its contribution to brain tumor risk. She contributed towards all the 100% of investigated areas.

Dr. Ayesha Saeed (AS) is a dedicated medical professional with a passion for the field of medicine. Holding a Doctor of Medicine degree from Dow University of Health Sciences, Pakistan. She has distinguished herself through exemplary clinical practice and a strong commitment to patient care and has a keen interest in improving patient outcomes.

**18. Aakash (AA)** is the co-author of the study. He contributed to the results and conclusions of the study, also contributed to the writing and editing of these sections. He also contributed to the references. He contributed to investigating how the brain inflammation induced disruption of gliogenic regulators (FGFR3, JAK-STAT, STAT3, S100, Hey1, HES1, DTX, IL-6, NF-κB, Neuregulin-1, MAPK/MEK, E2F/TCFL2, NFIX/Ephrins/Netrins) resulting in glial cell fate dysregulations and Its contribution to brain tumor risk. He contributed towards all the 100% of investigated areas.

Aakash MD, currently working in Florida State University Cape Coral Hospital and he is MBBS from Sindh Medical College - Dow University of Health Sciences, Karachi, Pakistan. He is passionate about pursuing fellowship in gastroenterology. His goal is also to translate emerging findings in research of disease development mechanisms/ origins into their clinical implications.

*The work and contributions of everyone have been described in detail, the order is randomized and the numbering is just for referencing purpose*.

## Abbreviations

FGFR3: Fibroblast Growth Factor Receptor 3
JAK-STAT: Janus Kinase-Signal Transducer and Activator of Transcription
STAT3: Signal Transducer and Activator of Transcription
S100: S100 Calcium Binding Protein
Hey1: Hairy and Enhancer of Split-1
HES1: Hairy Enhancer of Split 1
DTX: Deltex
IL-6: Interleukin 6
NF-κB: Nuclear Factor Kappa-Light-Chain-Enhancer of Activated B Cells
MAPK: Mitogen-Activated Protein Kinase
MEK: Mitogen-Activated Protein Kinase Kinase
Neuregulin-1: Neuregulin 1
E2F/TCFL2: E2F Transcription Factor/Transcription Factor 2
NFIX: Nuclear Factor I X
Ephrins: Eph Receptors and Their Ligands
Netrins: Netrin Family of Proteins
CNS: Central Nervous System
NSCs: Neural Stem Cells
NPCs: Neural Progenitor Cells
GFAP: Glial Fibrillary Acidic Protein
OLs: Oligodendrocytes
OPCs: Oligodendrocyte Precursor Cells
BMP: Bone Morphogenetic Protein
TLR: Toll-Like Receptor
TNF-α: Tumor Necrosis Factor Alpha
ROS: Reactive Oxygen Species
DNA: Deoxyribonucleic Acid
RNA: Ribonucleic Acid
mRNA: Messenger Ribonucleic Acid
BBB: Blood–Brain Barrier
RTKs: Receptor Tyrosine Kinases
PI3K: Phosphoinositide 3-Kinase
AKT: AKT Serine/Threonine Kinase
ERK: Extracellular Signal-Regulated Kinase
CREB: cAMP Response Element-Binding Protein
GSK3β: Glycogen Synthase Kinase 3 Beta
VEGF: Vascular Endothelial Growth Factor
EGFR: Epidermal Growth Factor Receptor
HIF-1α: Hypoxia-Inducible Factor 1 Alpha
PTEN: Phosphatase and Tensin Homolog

